# Interpreting epidemiological surveillance data: A modelling study based on Pune City

**DOI:** 10.1101/2024.09.13.24313615

**Authors:** Prathith Bhargav, Soumil Kelkar, Joy Merwin Monteiro, Philip Cherian

## Abstract

Routine epidemiological surveillance data represents one of the most continuous and current sources of data during the course of an epidemic. This data is used to calibrate epidemiological forecasting models, as well as for public health decision making such as imposition and lifting of lockdowns and quarantine measures. However, such data is generated during testing and contact tracing and not through randomized sampling. Furthermore, since the process of generating this data affects the epidemic trajectory itself – identification of infected persons might lead to them being quarantined, for instance – it is unclear how representative such data is of the actual epidemic itself. For example, will the observed rise in infections correspond well with the actual rise in infections? To answer such questions, we employ epidemiological simulations *not to study the effectiveness of different public health strategies in controlling the spread of the epidemic*, but to study the quality of the resulting surveillance data and derived metrics and their utility for decision making. Using the BharatSim simulation framework, we build an agent-based epidemiological model with a detailed representation of testing and contact tracing strategies based on those employed in Pune city during the COVID-19 pandemic, as well as to generate synthetic surveillance data. Infected persons are identified, quarantined and/or hospitalised based on these strategies. We perform extensive simulations to study the impact of different public health strategies and the availability of tests and contact tracing efficiencies on the resulting surveillance data as well as on the course of the epidemic. The fidelity of the resulting surveillance data in representing the real-time state of the epidemic and in decision-making is explored in the context of Pune city.

## 1 Introduction

Routine epidemiological surveillance data is generated by public and private entities as part of efforts to diagnose, treat and contain any outbreak^1^. For instance, during COVID-19 outbreaks in Pune city, daily surveillance data^2^ included (1) the number of tests conducted, (2) the number of individuals who tested positive and their demographic information, (3) the number of people hospitalised, (4) the number of deaths, and (5) contacts of identified positive cases. Such surveillance data has been used as input for several epidemiological forecasting models^3–9^ which have tried to estimate the course of the epidemic in India. Even though these forecasting models have their merits in assisting decision-making and policies, they come with their own set of challenges^10–12^. Similarly, surveillance data represents a noisy estimate of the actual epidemic and it is therefore important to understand its limitations. Some analytical results have been obtained to model the introduction of delays and under-reporting^13^, but it is unlikely that the complexities of the public health response itself, such as testing, contact tracing and quarantining as well and resource constraints (such as number of testing kits available) can be easily modelled in an analytical framework. Therefore, simulations appear to be a useful tool to help understand the relationship between surveillance data and the true epidemic.

In addition to forecast models, metrics derived from surveillance data such as Test Positivity Rate (TPR), Case Fatality Rate (CFR) and Reproduction Number (R_t_) have themselves also been used to inform public health interventions. The World Health Organization (WHO) advocated the use of TPR as a metric to indicate whether the epidemic is controlled^14^. While this recommendation was not prescriptive or data-driven, India, like many other countries, used TPR to gauge the true extent of the pandemic and subsequently implement public health measures^15^,^16^. All over India, different districts were demarcated into red, orange, or green zones^17^,^18^based on indicators derived from programmatic surveillance, including the number of daily cases and the extent of testing and surveillance. Moreover, local governments implemented strategies such as large-scale random testing to reduce the value of metrics like TPR^19^, hoping, in turn, to reduce the spread of the epidemic.

Even though such metrics provide useful insights into the nature of the true epidemic^20^, understanding the correspondence between the actual epidemic curve and such metrics alone remains a challenge. These indicators are often biased and may not pick up asymptomatic carriers of the disease or even symptomatic carriers who choose not to self-report due to social or economic reasons^21^. Moreover, practical issues like limited resources, accessibility, and errors in data collection or analysis can lead to undercounting^22^,^23^. Evidence of such undercounting has been reported through the use of serological surveys^24^,^25^. Thus, it is essential to understand the relationship between the true epidemic and the “observed” epidemic as inferred from surveillance data.

To this end, we build an agent-based epidemiological model using the BharatSim simulation framework^9^ that simulates an epidemic and the attendant public health response in the form of testing, quarantining, and contact tracing. We build this model in the context of the public health system response to the COVID-19 pandemic in the city of Pune, India. Several epidemiological models already exist in literature^26–31^ that simulate an epidemic by assigning different disease states to people throughout the course of the epidemic. We choose an agent-based approach since it allows us to specify characteristics such as geographical locations and activity schedules for each individual (See S1 Appendix). Likewise, this approach also allows us to track each individual’s disease state, testing and quarantining status, and identified contacts, throughout the pandemic. In addition to modelling the spread of the epidemic, we also consider counterfactual scenarios with different public health responses, thereby studying the relationship between derived metrics and the true epidemic.

### 1.1 COVID-19 in Pune city

Pune city is located in the state of Maharashtra in western peninsular India and has a current estimated population of 4.5 million. The smallest administrative units are the electoral wards or “prabhags”, and Pune consists of 41 prabhags, each containing approximately 100,000 people (Recent expansion of the city to include suburbs has increased this number. See https://www.pmc.gov.in/en/pmc-prabhag-rachna-2022). These prabhags are part of a larger administrative unit called a “ward”, and each ward has a health officer who makes operational decisions such as deploying personnel for contact tracing or disinfection.

The first case of COVID-19 in Pune city was reported on March 9, 2020. The city experienced three major waves of the pandemic – the first between May-September 2020, the second between February-May 2021 and the third between December 2021-January 2022^32^. Pune experienced a complete lockdown between March-June 2020 and wards with a high number of cases were quarantined from the rest of the city. Despite these strict containment measures, the rate of growth of cases continued to increase^33^ and very high prevalence was observed in an early serological survey^34^, suggesting that the spread of SARS-CoV2 within containment zones was fairly unrestricted. The Infection Fatality Rate computed using serological prevalence was comparable to results obtained elsewhere in the world^34^, suggesting that undercounting of COVID-19 related deaths in Pune city was not substantial. With improvements in treatment protocols, the case fatality rate in Pune declined almost monotonically between March 2020 and May 2021, though the burden of mortality was much higher in the second wave^35^.

Some compartmental epidemiological models were operationally deployed during the first and second waves and forecasts were used in infrastructure planning, especially for critical cases who required ventilator support^20^,^28^. While lockdowns were the main policy intervention used during the first year of the pandemic, a more fine-grained policy for restriction of movement and economic activity based on oxygenated bed occupancy levels and test positivity were employed from June 2021, after the end of the second wave^36–38^, with the explicit intention of reducing further spread or “breaking the chain”. Surveillance data was also used in estimates of prevalence and decision-making, using heuristic ideas relating case fatality rate, test positivity to actual prevalence, and allocation of limited testing kits^20^. The use of epidemiological surveillance data for decision-making is not unique to Maharashtra, and has been attempted elsewhere as well see, for example^39–42^. In the next section, we describe our epidemiological model and how the public health response was incorporated into it. As a first step, we do not attempt to incorporate interventions such as lockdowns to keep the model simpler and its results interpretable.

## 2 Materials and methods

### 2.1 Population Structure

We model a “Prabhag” of Pune city, consisting of 100,000 individuals. These individuals are distributed demographically based on the estimated population data for 2012-14^43^. Individual agents in our model can exist in one of 8 possible epidemiological “states”: (S)usceptible, (A)symptomatic, (P)resymptomatic, Mildly Infected (MI), Severely Infected (SI), (R)ecovered, (H)ospitalised, or (D)ead – representing the progress of the disease. The transition between different states is shown in Figure 1. A detailed description of the rates of transition is given below. We assume that recovered individuals cannot get reinfected. If all individuals were in a single location, i.e. if the population were well-mixed, this would correspond to an 8-compartment epidemiological model. See S2 Appendix for a detailed description of such a model. Our model incorporates a network structure wherein individuals “move” between different locations, based on a schedule defined as a function of their individual attributes such as their age, or disease state. For instance, an “employee” (any individual with an age under 60 years) travels daily to their office in the morning, spends one time-step (4 hours) in their neighbourhood, and comes back home in the evening. A schematic of our network is shown in Figure 1. Individuals whose place of work is a hospital are designated as health care workers. For a detailed description of the schedules, see S1 Appendix. We model an isolated Prabhag with no movement of the infection across Prabhag boundaries.

**Figure 1.**
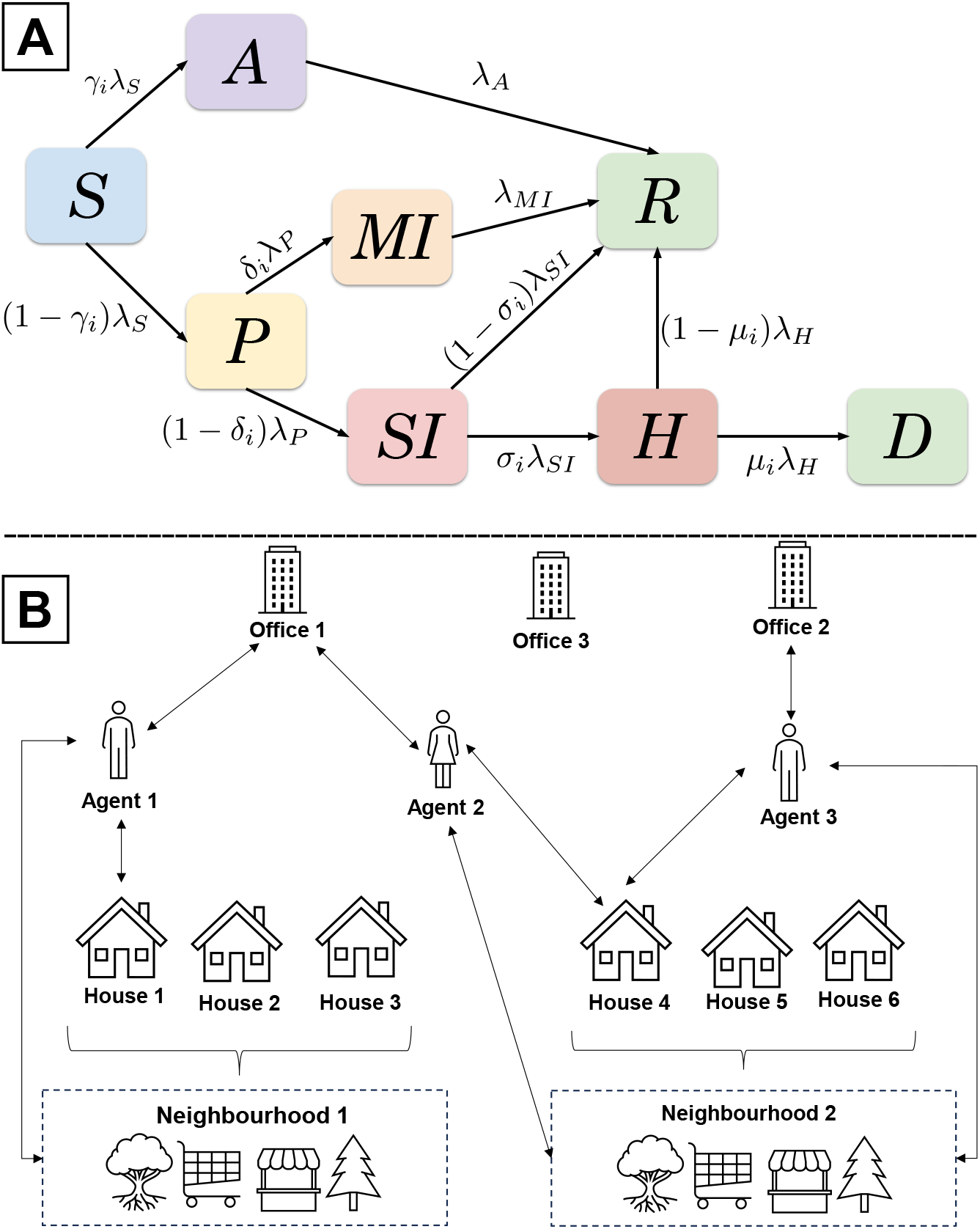
(A) A schematic describing the eight disease states and the transitions between them. (B) A schematic of the geographical structure depicting the movement of people between different locations. The agents (individuals) depicted here are employees who travel daily to their office, spend one time-step in their neighbourhood and come back home.

### 2.2 Simulating the Epidemic

The epidemic is seeded by randomly choosing a set of people and infecting them. The number and location of such seed individuals can be varied to model different scenarios. For example, infecting a large fraction of employees at a particular office mimics a super-spreader event. In our experiments, we choose 100 random seed individuals (on average) in a locationindependent manner and set their disease state to Asymptomatic at the start of the simulation. Each network location in our model is considered to be well-mixed. The number of individuals in each of these locations varies with time as people move between their assigned homes, workplaces, and neighbourhoods. As shown in Figure 1, individuals transition stochastically, at specified rates, from their current disease state to other accessible states. To shuttle individuals between compartments, we define a variable, *λ*_*DS*_, (where DS is the disease state), which is the rate at which an agent exits that state, set by considering the reciprocal of the average time spent in that disease state. For instance, *λ*_*A*_ is the constant rate at which an agent exits the Asymptomatic state. The probability of exiting a disease state in an interval Δ*t* is just *λ*_*DS*_Δ*t*. For states that branch into two states, we further define a branching probability – *γ, µ, δ* and *σ* in Figure 1A – as the probability that an individual enters one of the two branched states post exiting the primary state. The subscript *i* is added since these variables are age-stratified. For example, for a given age group, *γ*_*i*_ is the probability that an agent becomes Asymptomatic after exiting the Susceptible state. Similarly, 1*− γ*_*i*_ is the probability that the agent becomes Presymptomatic. The values assigned to all variables are given in Table 1, along with a short description of what they mean.

**Table 1.** (top) Values assigned to the variables of the form *λ*_*DS*_, where DS stands for disease state. The values have been adopted from^26^ and references therein. For a Markov state model, the reciprocal of these values corresponds to the average number of days spent in that disease compartment. A comparison of the epidemic curves for different values of *λ*_*S*_ and their R_0_ values is presented in S2 Appendix. (bottom) Age-stratified values for *γ, µ, δ* and *σ* . ^⋆^Values modified from the original reference to ensure that roughly 60-70% of the population is recovered at the end of the epidemic.

| Parameter | Value (day<sup>-1</sup>) | Description |
| --- | --- | --- |
| $\lambda_S$ | 0.5 | Rate at which infected individuals can infect the susceptible population. Corresponds to $R_0 = 1.5$. |
| $\lambda_P$ | 0.5 | Individuals remain Presymptomatic for an average of $1/\lambda_P = 2$ days. |
| $\lambda_A$ | 0.143 | Asymptomatic individuals recover in an average of $1/\lambda_A = 7$ days. |
| $\lambda_{MI}$ | 0.1 | Individuals with mild symptoms recover in an average of $1/\lambda_{MI} = 10$ days. |
| $\lambda_{SI}$ | 0.143 | Individuals with severe symptoms get hospitalised or recover in an average of $1/\lambda_{SI} = 7$ days. |
| $\lambda_H$ | 0.143 | Individuals who are hospitalised recover or die in an average of $1/\lambda_H = 7$ days. |

**Table 1.** (top) Values assigned to the variables of the form $\lambda_{DS}$ , where DS stands for disease state.
| Age group | 0-9 | 10-19 | 20-29 | 30-39 | 40-49 | 50-59 | 60-69 | 70-79 | 80-89 | 90+ |
| --- | --- | --- | --- | --- | --- | --- | --- | --- | --- | --- |
| Asymptomatic probability – $\gamma^{*28}$ | 0.50 | 0.45 | 0.40 | 0.35 | 0.30 | 0.25 | 0.20 | 0.15 | 0.10 | 0.10 |
| Death probability – $\mu^{*28}$ | 0.0555 | 0.0561 | 0.0429 | 0.0498 | 0.1020 | 0.1500 | 0.2910 | 0.6300 | 0.6600 | 0.6600 |
| Mild Infection probability – $\delta^{*28}$ | 0.999 | 0.970 | 0.920 | 0.880 | 0.840 | 0.800 | 0.760 | 0.720 | 0.680 | 0.640 |
| Hospitalization probability – $\sigma^{*44}$ | 0.0088 | 0.0240 | 0.0630 | 0.1700 | 0.4600 | 1.0000 | 1.0000 | 1.0000 | 1.0000 | 1.0000 |

With the exception of the Susceptible state, all transition rates out of the different states are constant in time. To find the rate of infection of a susceptible agent, we first compute the effective infected fraction of individuals at the location of that agent, given by:

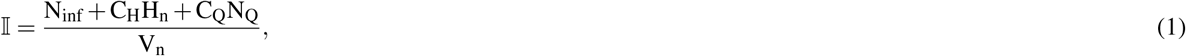

where N_inf_ is the total number of infected people in a given location who are not isolated or quarantined, H_n_ is the number of hospitalised people (which is only non-zero if the location is a hospital), and N_Q_ is the number of infected people who are quarantined. This includes people who have tested positive and are quarantined, asymptomatic low-risk contacts, symptomatic people who are isolated before getting a test and tested people waiting for their test result (see subsection 2.3 below). As agents are always isolated or quarantined only in their households, this variable is only non-zero for household locations. C_H_ is a scaling factor that decreases the transmissivity of a hospitalised person. We incorporate this factor to take into account the effect of protective measures like PPE kits enforced by hospitals that reduce the infectivity of patients. In our model, C_H_ is set to 0.1^26^. Similarly, C_Q_ is a scaling factor which reduces the transmissivity of quarantined and isolated agents. In this work, we also set C_Q_ = 0.1^26^. This is meant to represent an idealized scenario in which quarantine is effective in reducing probability of disease transmission by 90%. In S6 Appendix, we repeat our simulations with different values of *C*_*Q*_, and show that varying this parameter does not produce qualitatively different results (Figure S10).

V_n_ is the total number of people at that location defined as

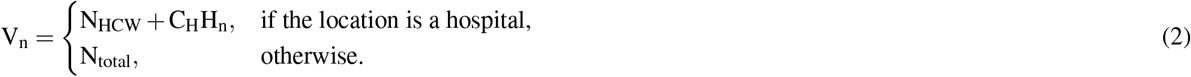

Here, N_HCW_ is the number of health care workers in the hospital, and N_total_ is the total number of individuals in that location. The exposure probability 𝔼 is given by

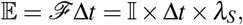

where Δ*t* is the duration of one time-step, *λ*_*S*_ is the transmission rate, and *ℱ* = *λ*_*S*_× 𝕀 is the force of infection. This is the probability that an individual exits the Susceptible state, i.e., gets infected. The exposure probability is calculated for every susceptible individual at each time-step at any given location, according to their pre-defined schedules.

### 2.3 Modelling the Public Health Response

Our model for the public health response is based on the strategy adopted at the Pune city COVID-19 “war-room”, a central data-gathering hub during 2020-2022, where one of the authors (JMM) worked as a volunteer during 2020-21. We elicited information regarding strategies to identify individuals for testing, contact tracing and quarantine. Analysis of contact tracing data between March-June 2020 (see S5 Appendix) suggested that on average, around seven contacts were identified for each index patient and contacts were designated as high or low-risk based on proximity and duration of contact (see following sections for more details).

While introducing testing, we consider scenarios where the start of the actual epidemic is not coincident with the start of the public health response. For instance, in India, while the first case of COVID-19 was detected in January 2020^45^, the availability of tests was limited until May 2020, when about 453 tests per million people became available^46^. To mimic this scenario, we activate testing only after a threshold number of people “self-report” themselves as being sick. Unless stated otherwise, this testing threshold is set at 1% of the population for our experiments.

There are three possible interventions through which an individual becomes eligible for testing – Self-Reporting, Contact Tracing, and Random Testing. Depending on the public health response, some of these interventions may or may not be active. We assume that there are limited tests every day and thus, not everyone who becomes eligible for getting a test is immediately tested. Depending on the number of tests, individuals are randomly selected from the list of all eligible people and are tested based on a priority order described below. Note that individuals who are eligible for getting tested are isolated until they receive their test.

#### Self-reporting

Individuals exhibiting symptoms of COVID-19 can self-report and become eligible for receiving a test. Similar to real life, not all individuals will report their symptoms, and there is a probability associated with reporting symptoms. People with severe symptoms are more likely to self-report than people with mild symptoms. In all our experiments, severely infected people self-report with a probability of 0.8, while mildly infected people self-report with a probability of 0.4. Moreover, not all people exhibiting symptoms are infected with COVID-19. During the epidemic, other diseases like the flu are still prevalent, and people infected with them exhibit similar symptoms^47^. To mimic this in our model, a small fraction of individuals (on average, 0.025% of the susceptible population) infected with flu-like illnesses (but still susceptible to COVID-19) report their symptoms every day and become eligible for getting a test. For simplicity, we assume that the people infected with or recovered from COVID-19 are not susceptible to other flu-like illnesses.

#### Contact tracing

For every agent who tests positive, we designate a list of “contacts” to represent those individuals who might have interacted with that agent. All individuals from the same household and a fraction of individuals belonging to the same office and the same neighbourhood as the agent who tested positive are selected as contacts. All household contacts (irrespective of their symptom status) and all symptomatic office and neighbourhood contacts are classified as high-risk contacts, and are made eligible for testing. All the asymptomatic office and neighbourhood contacts are classified as low-risk contacts, and they are isolated for 7 days.

#### Random testing

Individuals are randomly selected from the population and are made eligible for getting a test. These individuals are selected only if they are not already hospitalised or eligible for a test through any other interventions mentioned above. Moreover, individuals who have tested positive, quarantined, or isolated are also not selected.

#### Priority order for testing

At the peak of the first wave of the epidemic, only about 12,000 tests were conducted daily in the Pune district, which is home to approximately 12 million people^48^. National statistics also paint a similar picture^49^, and about a million tests were performed daily for a population of around 1.4 billion people. Considering the limited number of tests, the Indian Council of Medical Research formulated a set of guidelines to test individuals based on a priority order^50^. We simulate such guidelines with a limited number of daily tests and a priority order for who receives the test. The priority order is as follows:

1. Self-reported symptomatic individuals and their high-risk contacts,

2. People eligible via random testing.

Since there are a fixed number of daily tests, the people eligible for a test are pooled together based on the priority order. Tests are distributed once a day, during which time people are randomly sampled from this pool and given a test. For instance, consider the Self Reported + Random Testing public health response scenario – self-reported symptomatic individuals are collected in pool 1, and those eligible for random testing are collected in pool 2. According to the priority order, people are first randomly sampled from pool 1 and given a test, and if (and only if) any tests are remaining are people then randomly sampled from pool 2 and tested. We assume all our tests to be RT-PCR tests, which have a 100% specificity and 100% sensitivity, thereby ensuring no false positives or false negatives. To account for the time delay in declaring results of RT-PCR tests^51^, we introduce a two-day delay between an individual getting tested and receiving the test result. All positively tested individuals are quarantined in their homes for 14 days to account for the incubation period of COVID-19^52^.

#### Calculation of Case Fatality Rate (CFR)

We compute the cumulative case fatality rate (CFR) by dividing the cumulative number of deaths by the cumulative number of identified infections. The number of identified infections includes the number of individuals identified using testing and those who are dead. This is inline with a policy issued by the Indian Council for Medical Research (ICMR) during the first wave of the pandemic in India^53^. Note that in our simulations we only account for deaths from COVID-19.

#### Calculation of R_t_

R_t_, also known as the effective reproduction number, is an estimator of the number of infections caused by one infected person. During the COVID-19 pandemic, public health systems used R_t_ to guide and direct public health responses^54^.

We compute R_t_ from the underlying simulation data by using its definition as the number of secondary infections due to a new primary infection. In our model, susceptible agents are infected through a force of infection that depends on the total number of infected agents in their location at that time-step. Once an agent is infected, we assign them an “infector”, drawn randomly from all the infected agents present at that location in the previous time-step. The underlying assumption in this approach is that all infected individuals at a location have an equal probability of transmitting the infection to a susceptible agent. This method can be extended to introducing individual-level heterogeneity in infectivity, but is beyond the scope of this work. In this way, we record the number of secondary infections caused by every infected agent. From this, we compute R_t_ as being the average number of secondary infections of caused by an agent who was infected on day *t*.

To estimate R_t_ from public health data, we use the R Package, EpiEstim^55^. EpiEstim computes R_t_ using only daily incidence data and the serial interval distribution – the estimated time between symptom onset in a case and their infector. This quantity is called EpiEstim R_t_. EpiEstim has been validated against both simulation and public-health data for COVID-19^55^. We use the “parametric si” method with a mean serial interval of 6.3 and a standard deviation of 4.2^56^ to calculate R_t_ on weekly sliding intervals. We use previously determined serial interval values since the actual values are not known a priori. We check the validity of these values by comparing the EpiEstim R_t_ calculated through true infections (instead of the identified infections) and the real R_t_ (see Figure S13 in S7 Appendix).

#### Ethics Statement – Analysis of Contact Tracing Data from Pune

Anonymized contact tracing data was used to compute the average number of contacts. The contact tracing data was derived from programmatic data without personal identifiers, hence individual patient consent was not obtained as infeasible. The Ethics Committee of Indian Institute of Science Education and Research, Pune, India approved the analysis of COVID-19 programmatic data and has waived the need for obtaining the consent. The analysis and reporting were performed in accordance with the relevant guidelines and regulations.

### 2.4 Experimental Design

A public health response is a combination of different interventions that can be implemented. The interventions are (a) **Self Reported (SR)**: Only people who self-report are tested, (b) **Contact Tracing (CT)**: Identified high-risk contacts are tested, and low-risk contacts are isolated and (c) **Random Testing (RT)**: people are randomly sampled from the population and tested.

To compare the relationship between metrics and the true state of the epidemic, we conduct four sets of experiments:

1. Variation of *λ*_*S*_ in the absence of any public health intervention, where *λ*_*S*_ is the infection rate.
2. Variation of the public health response given a fixed number of 500 daily tests. We use the following combinations of interventions
  a. Self Reported (SR)
  b. Self Reported along with Contact Tracing (SR+CT)
  c. Self Reported along with Random Testing (SR+RT)
  d. Self Reported along with Contact Tracing and Random Testing (SR+RT+CT)
3. Variation of the number of daily tests given a fixed public health response (SR+RT+CT)
4. Variation of the efficiency of contact tracing given a fixed number of 500 daily tests and a fixed public health response - SR + RT + CT. This involves varying two parameters, *f*_*O*_ and *f*_*N*_ which describe the fraction of identified contacts at the office and neighbourhood respectively. All the values of the described parameters which are varied during each experiment are given in Table 2. Since our model is inherently stochastic in nature, we ran 30 simulations for each set of parameters. We present the mean values of all our computed quantities as well as the 1*σ* standard deviations.

**Table 2.** Description of the parameters varied during each experiment. There are four different public health response scenarios - (1) **SR** Only people who self-report are tested, (2) **SR + CT** People who self-report, identified high-risk contacts and low-risk symptomatic contacts are tested, (3) **SR + RT** People who self-report and people who are randomly tested, (4) **SR + RT + CT** People who self-report, identified high-risk contacts, low-risk symptomatic contacts as well as people who are randomly sampled from the population are tested. Note that there are 40 employees in an office and 400 people in a neighbourhood on average (see **Supplementary Material Section S1.2** so the following fractions translate to 2(10), 6(30), and 10(50) contacts identified at the office (neighbourhood) on average per individual. **SR** - Self Reported, **CT** - Contact Tracing, **RT** - Random Testing

| Sr.no | Description | Variation of parameters |
| --- | --- | --- |
| 1 | Variation of $\lambda_S$ | 0.3, 0.4, 0.5 |
| 2 | Variation of public health scenarios given 500 daily tests | Self Reported<br>Self Reported + Contact Tracing<br>Self Reported + Random Testing<br>Self Reported + Contact Tracing + Random Testing |
| 3 | Variation of the number of daily tests given a fixed public health response - Self Reported + Contact Tracing + Random Testing | 100, 200, 300, 500, 700 |
| 4 | Variation of efficiency of contact tracing (fraction of contacts identified at office, fraction of contacts identified in the neighborhood) given 500 daily tests and a fixed public health response - Self Reported + Contact Tracing + Random Testing | **low** - (0.05, 0.025)<br>**medium** - (0.15, 0.075)<br>**high** - (0.25, 0.125) |

## 3 Results

### 3.1 Infection curves for different transmission rates

We ran simulations for three different values of the transmission rate, *λ*_*S*_ – 0.3, 0.4, and 0.5 –in the absence of any public health interventions. A comparison of the infection curves for these values is presented in Figure S2 in S2 Appendix. Upon analysing the results of these simulations, we chose *λ*_*S*_ = 0.5 for all our further experiments to ensure that around 60 *−* 70% of the population is infected by the end of the epidemic. From our calculations of R_0_, described in S2 Appendix, we find that simulations with this *λ*_*S*_ correspond to an *R*_0_ of 1.5. The data used to calibrate *λ*_*S*_ to *R*_0_ is provided in Figure S1. We do not analyse these experiments further since it is not in the scope of the current work.

### 3.2 Effects of different public health response scenarios

We model different public health response scenarios given a fixed number of 500 daily tests. We choose 500 daily tests since this was similar to the maximum number of daily tests (per 100,000 people) that were available in Pune during the second wave (Feb-May 2021)^48^. For the scenarios when contact tracing is active, we set *f*_*O*_ and *f*_*N*_ to be equal to 0.1 and 0.02 respectively which means that on average, 4 colleagues and 8 neighbours are identified as contacts of each positively tested person.

The left column of Figure 2 shows the evolution of the Test Positivity Rate (TPR) across different public health response scenarios. It is evident that contact tracing effectively reduces the Test Positivity Rate (TPR). Whenever the contact tracing intervention is active, self-reported symptomatic individuals, high-risk contacts, and low-risk symptomatic contacts have an equal priority to get tested. Since the number of daily tests is fixed, a significant fraction of the tests are used on high-risk contacts who are not necessarily infected, and this brings down the TPR. Random testing has no effect on the TPR in the initial days of the epidemic. This is because the number of daily tests available is fixed and given the priority order, there are barely any tests left for random testing. If the number of daily tests is increased, a larger fraction of tests will be used for random testing (see Figure S15 in S8 Appendix) which will lead to a reduction in the TPR. At the end of the epidemic, we see a secondary peak in TPR in the absence of random testing. At this time, only a small fraction of individuals remain infected and since infection by COVID-19 precludes infection by the flu, the probability that a symptomatic individual is COVID-19 positive increases significantly.

**Figure 2.**
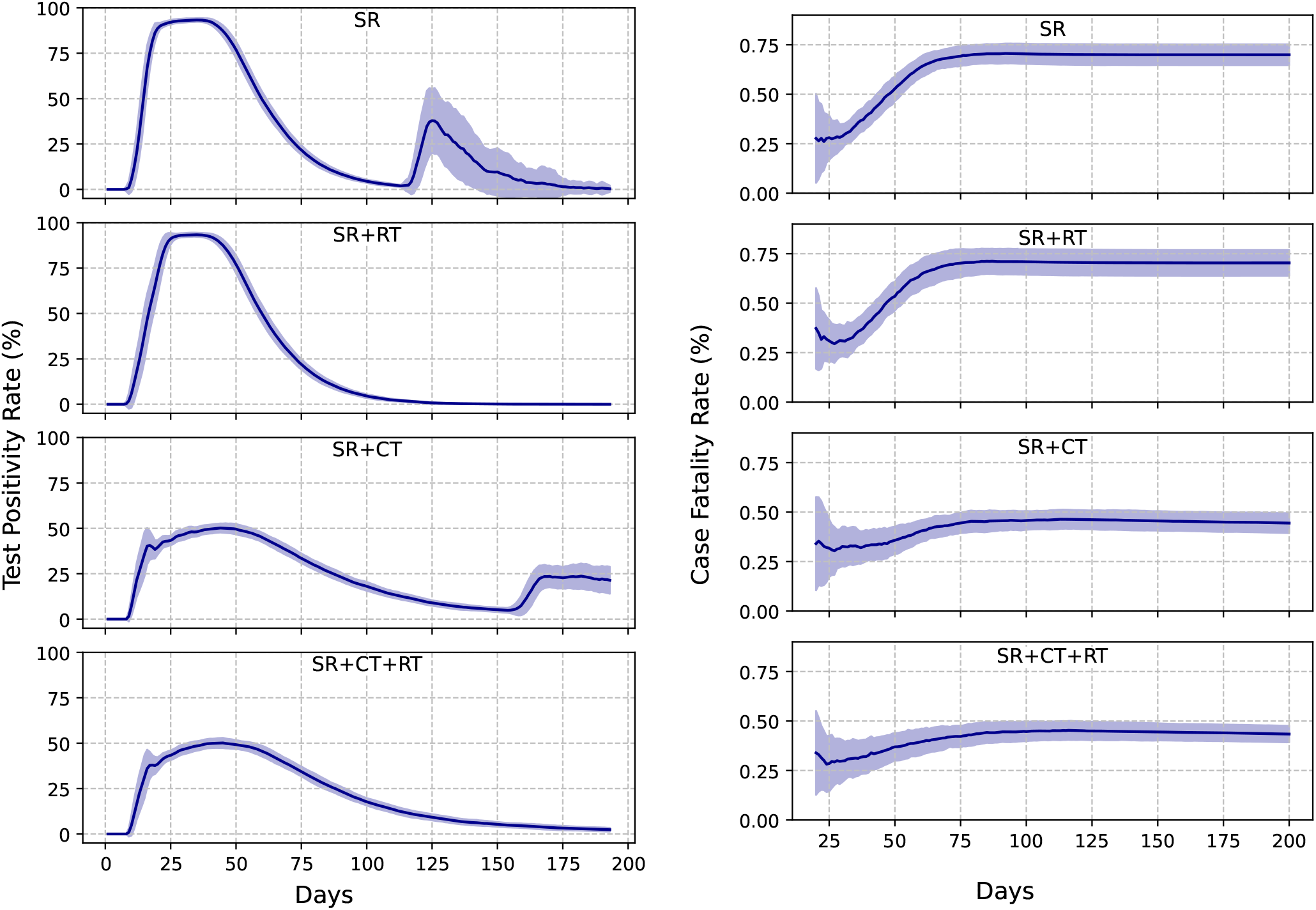
The left and right panels show the evolution of the Test Positivity Rate (TPR) and the Case Fatality Rate (CFR) respectively, for different public health response scenarios given a fixed number of 500 daily tests. The solid lines depict an average of the metrics taken over 30 individual simulations and the shaded regions denote the 1*σ* spread. Furthermore, each metric curve is smoothed over using a 7-day rolling average.

This effect also increases the 1*σ* spread in TPR as the number of symptomatic people getting tested towards the end of the epidemic is highly stochastic. This result suggests that increases in TPR after an observed epidemic peak declines^19^ may be simply due to this reason, and not indicative of a new wave of infections. When random testing is active, a majority of tests are used for random testing at the end of the epidemic, reducing TPR and mitigating this effect, as shown in the respective panels in Figure 2. However, we note that this reduction in TPR is purely “cosmetic” in the sense that it has no impact on health outcomes (see Figure S3 in S3 Appendix). Note that our TPR values are significantly higher as compared to actual reported values in Pune city (see Figure S9 in S5 Appendix). This is because we are only analyzing a population contained within a single hypothetical ward. There is no migration of people between different wards and hence, it is easier to identify and test people with symptoms as well as trace contacts. The right panel of Figure 2 shows that contact tracing effectively reduces the cumulative Case Fatality Rate (CFR).

The left column of Figure 3 compares the evolution between the actual number of infected people (or the number of true infections) and TPR. There is an indication of a weak phase relationship as both of these curves rise and fall together. The second peak in TPR for the SR and SR + CT scenarios is due to the absence of random testing as mentioned above. The right panel of Figure 3 compares the evolution of the rate of change of true infections with the rate of change of TPR. The stars denote the corresponding days on which this rate goes to 0 or the days on which the infection or TPR curves peak. The TPR peaks slightly before the infection for all the public health response scenarios considered. Moreover, for all scenarios, the rate of change of TPR always peaks before the rate of change of the number of true infections and subsequently the true infection peak (given by the location of the red star). These phase relationships remain robust even under public health scenarios where quarantining is less effective in limiting transmission (see Figure S10). This result suggests that the rate of change in TPR is a promising indicator for the peak of the actual number of infections. In the absence of random testing, the rate of change of TPR shows a small peak at the end of the epidemic which is a manifestation of the second peak in TPR.

**Figure 3.**
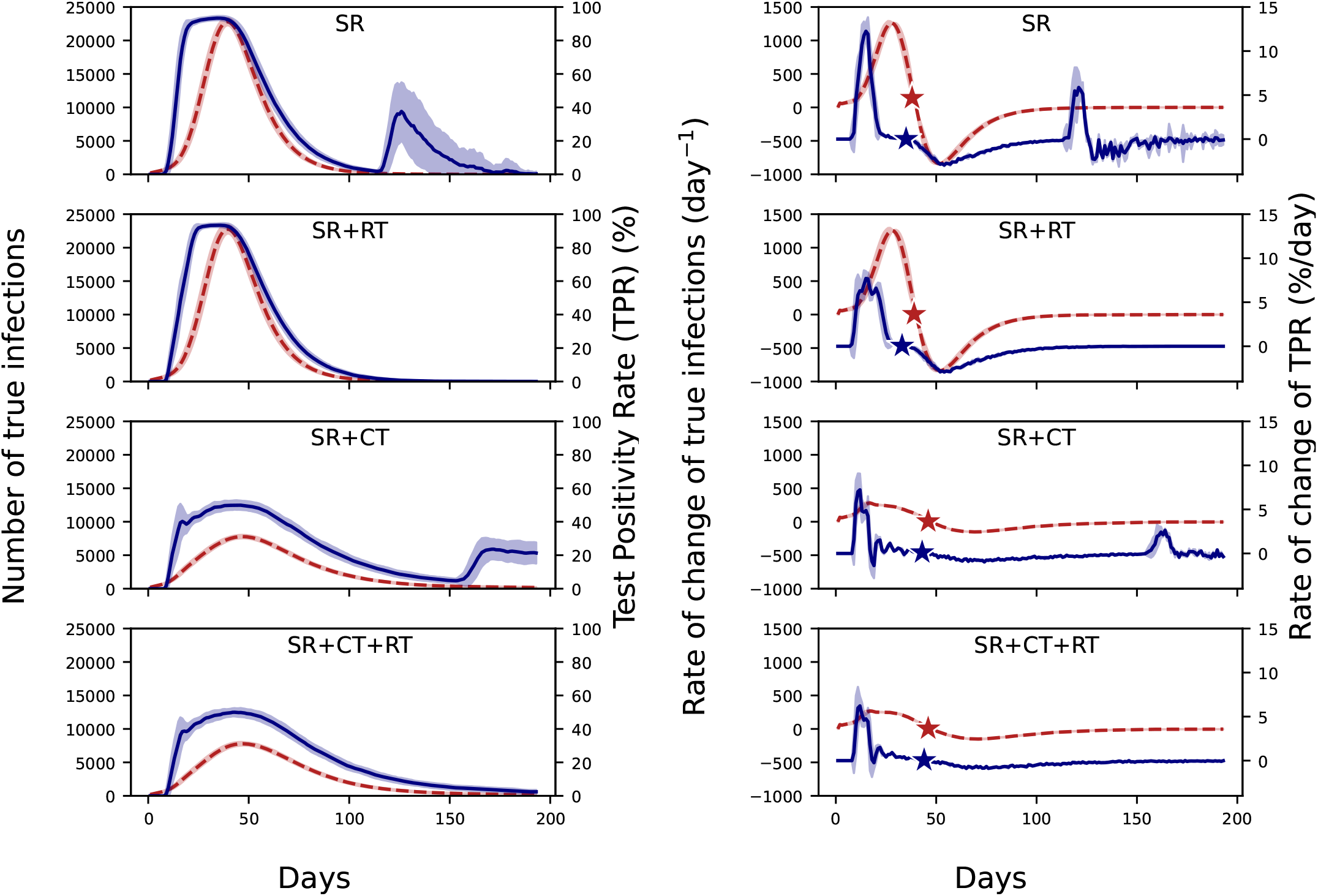
The left panel shows a comparison between the metric TPR (solid blue line) with the number of true infections (dashed red line) for different public health responses. The right panel shows a comparison between the rate of change of metric TPR (solid blue line) with the rate of change of the number of true infections (dashed red line) for different public health responses. The stars denote the days on which the rate of change of true infections or the metric TPR goes to 0 or in other words, the days on which the infection and the TPR curves peak. All the plots correspond to a 7-day rolling average.

A similar analysis comparing the evolution of the number of true infections and the Case Fatality Rate (CFR) is shown in Figure S7 in S4 Appendix. For all the public health response scenarios, the cumulative CFR asymptotes to a constant value after the peak of the infection curve and thus, CFR does not give prior indication of the peak of the infection curve. However, the peak of the infection curve occurs between the peak of the rate of change of TPR and the time when the cumulative CFR flattens out. Thus, these two metrics together can be used as indicators for the peak of the epidemic being reached.

### 3.3 Effects of the daily testing rate

In this experiment, we vary the number of daily tests given a fixed public health response (SR + RT + CT). *f*_*O*_ and *f*_*N*_, the fraction of identified contacts at the office and neighbourhood, are set to 0.1 and 0.02 respectively. Figure S4 in S3 Appendix shows the evolution of the number of true infections and the number of cumulative deaths for different numbers of daily tests. It is observed that the number of true infections and the number of cumulative deaths decrease with an increase in the number of daily tests. However this decrease is not linear as both these quantities saturate after a subsequent increase in the number of daily tests. Figure 4A depicts the evolution of the Test Positivity Rate (TPR) and the Case Fatality Rate (CFR). Both these metrics show a reduction as the number of daily tests are increased. We attribute the reduction in the TPR to the fact that as the number of tests are increased, the fraction of positive tests reduces while the reduction in CFR is a consequence of the reduction in the number of deaths. However, this reduction is non-linear in both the cases. Even after a 7-fold increase in the number of tests from 100 to 700, the peak of the TPR only reduces roughly by a factor of half, suggesting that increasing testing with the aim to reduce peak TPR may not be very effective. As the number of tests increase 7-fold, the peak of the TPR curve drops by ∼ 40% and the peak of the infection curve also drops by roughly the same amount. However, the CFR drops by ∼ 90% while the number of cumulative deaths reduces by ∼ 25%. Thus, our simulations indicate that the change in TPR could be a reliable indicator of the change in the number of true infections whereas change in CFR is not a reliable indicator of likely changes in mortality.

**Figure 4.**
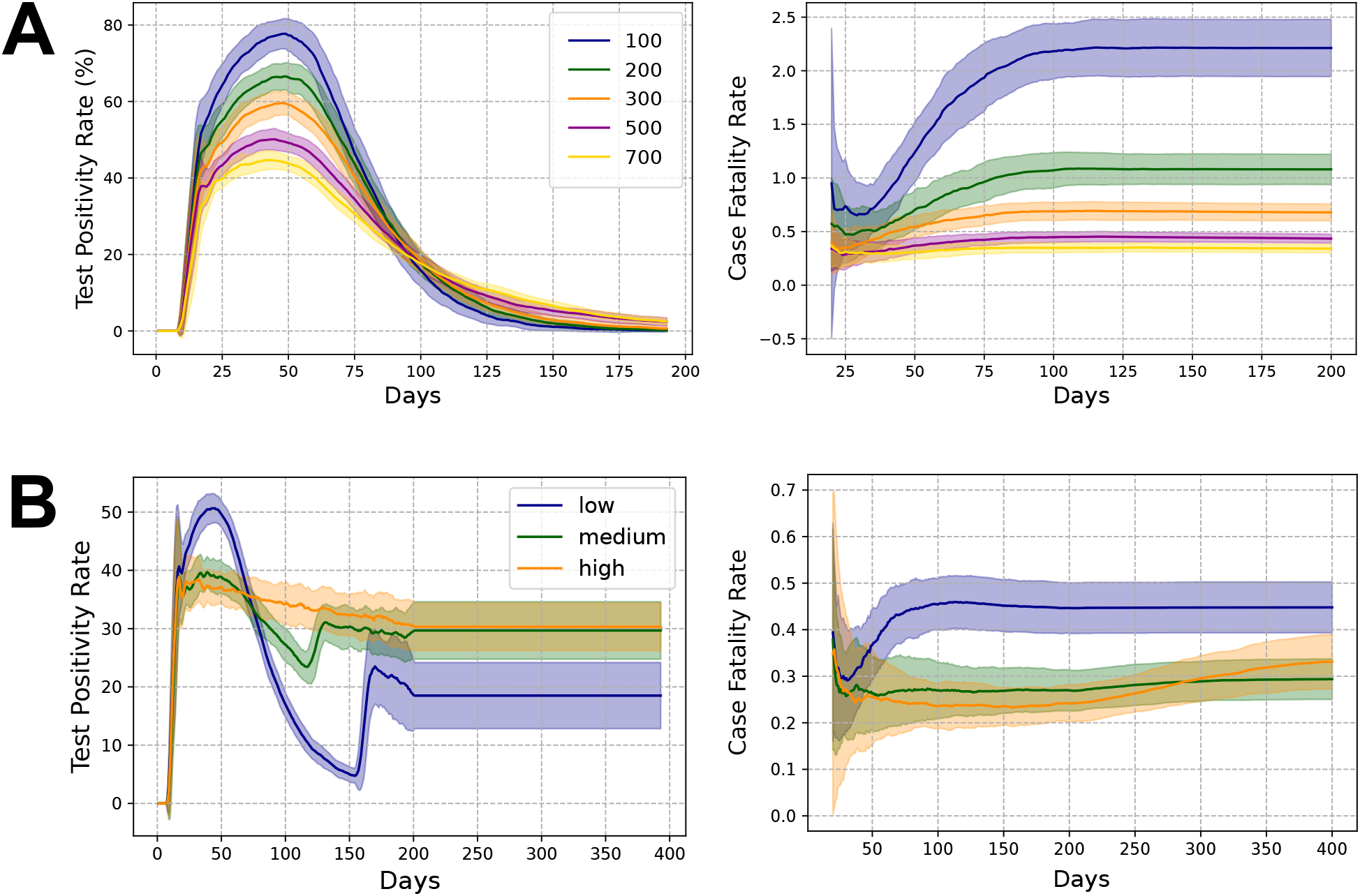
Evolution of the Test Positivity Rate (left) and the Case Fatality Rate (right) for (A) different number of available daily tests, given a fixed public health response - SR + CT + RT, and (B) different efficiencies of contact tracing (low, medium, and high), given a fixed public health response - SR + CT and a fixed number of 500 daily tests. All plots correspond to a 7-day rolling average.

We show the phase relationship curves between TPR and the number of true infections for the rest of the experiments in S6 Appendix. The left panel of Figure S11 shows that we observe a consistent phase relationship between TPR and the number of true infections. As shown before, the peak of the rate of change of TPR is seen slightly before the peak of rate of change of number of true infections and much before the peak of the infection curve itself (given by the location of the red star). This again indicates that the rate of change of TPR could be a reliable metric for forecasting the peak in the number of true infections. As observed earlier, the asymptotic behaviour of the CFR curve occurs after the peak of the number of true infections (see Figure S8 in S4 Appendix).

### 3.4 Effects of different efficiencies of contact tracing

In this section, we explore the isolated effects of the “efficiency” of contact tracing. We simulate a fixed public health response - SR + CT and vary the values of *f*_*O*_ and *f*_*N*_ - (i) (0.05, 0.025), (ii) (0.15, 0.075), and (iii) (0.25, 0.125), given a fixed number of 500 daily tests. These respective scenarios are labelled as low, medium and high and these labels characterize the efficiency of contact tracing and thus, the number of contacts identified for each positively tested person. Figure S5 in S3 Appendix shows the evolution of the number of true infections and the number of cumulative deaths for all three contact tracing efficiencies. Since the infection wave does not end even after 200 days, we extend our simulation time to 400 days.

The left column of Figure 4B shows that increasing the efficiency of contact tracing reduces the peak of the TPR, but increases the spread of the TPR curve. For example, when the contact tracing efficiency is high, TPR shows a gradual decline and only reduces to about 75% of its maximum value even after 400 days. The reasons for this are twofold; (i) the length of the epidemic itself is extended, and (ii) increasing the efficiency increases the number of identified contacts and a significant fraction of tests are utilized on testing them, leaving a constant symptomatic pool of people within the testing queue who will get tested positive. Thus, in this case, TPR does not capture the dynamics of the epidemic as the TPR curve reflects the positivity of those in the queue rather than the current state of the epidemic curve. The second peak in TPR is due to the absence of random testing, as explained above.

Figure 4B shows that the reduction in CFR and TPR is not linear with an increase in the efficiency of contact tracing and both metrics exhibits a threshold-like behaviour. This is evident from the fact that increasing the contact tracing efficiency from medium to high has negligible effect on either CFR or TPR. However, the health outcomes do not show a similar trend and are markedly different. As the contact tracing efficiency is increased from low to high, the peak of the infection curve drops by ∼ 70% and the total number of deaths drops by ∼ 35%. The metrics do not show a proportionate reduction as the TPR only drops by ∼ 20% and the CFR drops by ∼50%. Thus it is difficult to draw any inferences about the state of the epidemic and health outcomes just from changes in CFR and TPR when the efficiency of contact tracing changes. Furthermore, our results suggest that a high sustained value of TPR may not indicate a suboptimal public health response – on the contrary, it suggests that in resource constrained settings, a high sustained TPR may in fact indicate that contact tracing is being carried out in a more effective manner. Conversely, a rapid drop in TPR (as observed in the low-efficiency CT scenario) is not necessarily indicative of better health outcomes over the course of the epidemic wave.

Our analysis on the phase relationships yield similar results as earlier (see Figure S12); the peak of the rate of change of TPR happens much before the peak of the epidemic given by the location of the red star, and thus the rate of change of TPR can forecast the peak of the epidemic.

### 3.5 Relationship between undetected cases and CFR

Estimating the number of undetected cases is important to make decisions regarding intensifying the levels of testing or changing the testing strategy. In the case where the infection fatality rate (IFR) is known, a straightforward estimate of undetected cases is provided by the CFR simply as the ratio between the CFR and IFR. However, under-reporting as well as the lag between the onset of infection and eventual death (i.e, survival times) can complicate this relationship, and efforts have been made to develop estimators of CFR for such scenarios^57^. Here, we use our simulations to explore the relationship between CFR and undetected cases at different times during the epidemic and also look at its sensitivity to the different public health scenarios that we have simulated.

Figure 5 shows the relationship between undetected cases and CFR for all our simulations, with each individual run from each experiment represented by a single point. One clear conclusion is that a small number of tests (the 100 daily tests – DT 100 – scenario) can lead to a large range of CFRs for the same fraction of undetected cases, even when contact tracing is active. This is true for all times of the epidemic, even when nearing the end of the wave when the CFR has become relatively constant (see Figure 4A).

**Figure 5.**
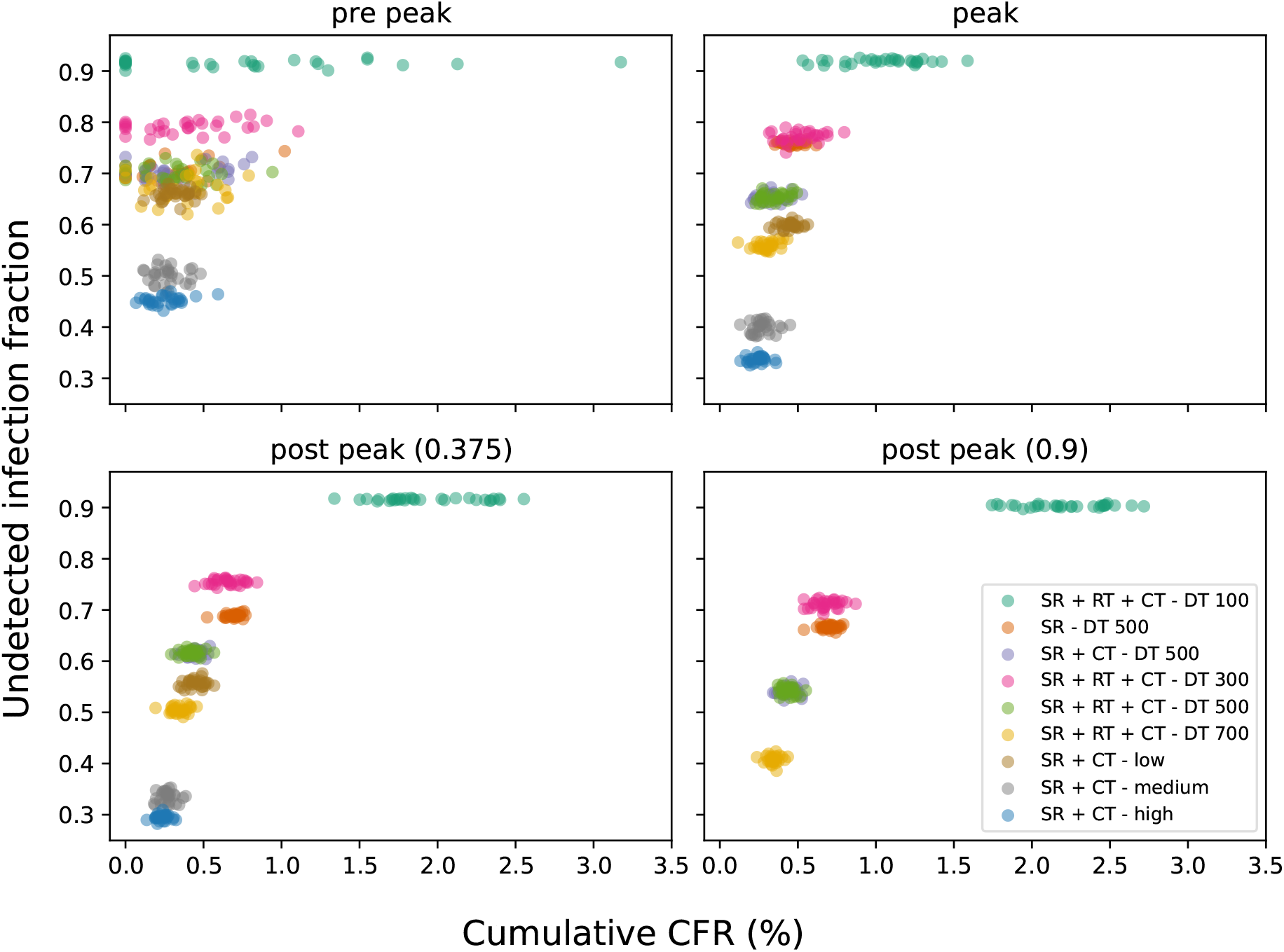
Scatter plot of the fraction of undetected cases against the cumulative CFR for different public health response scenarios.Panels correspond to different time points in the epidemic: pre-peak, peak, and post-peak (with post-peak panels labeled by the fraction of the simulation time (200 days or 400 days)). Each dot represents a single run and each colour represents a single scenario. Two of the simulations for the DT 100 case resulted in extremely high CFR values and were excluded from this plot for the purpose of better presentation.

In most scenarios, CFR provides a poor estimate of undetected cases until the peak of the epidemic, even though this is the period when public health decision making is crucial. Only near the end of the epidemic is there a reasonable relationship between CFR and the fraction of undetected cases (excluding the DT 100 scenario).

However, even for this purpose, including contact tracing (even at low efficiency levels) along with a higher number of tests results in the least spread in the relationship between CFR and fraction of undetected cases at all times of the epidemic.

### 3.6 Fidelity of estimated R_t_

Effective Reproduction Number (R_t_) is a time-varying quantity which represents the average number of secondary cases for every primary case at time *t*. An accurate estimate of R_t_ is important to assess effectiveness of interventions and is commonly used to judge the state of the epidemic and guide public health response. To examine the fidelity of R_t_ estimated using surveillance data in capturing the growth and decay phase of the epidemic, we plot R_t_ estimated using EpiEstim^55^ against the actual value of R_t_ computed from our simulations. This comparison allows us to evaluate the impact of data generated by different public health response scenarios in the estimation of R_t_.

For all our experiments, we observe that the estimated value of R_t_ deviates from the actual R_t_ during the initial part of the epidemic(Figure 6). Furthermore, EpiEstim’s confidence in its R_t_ estimates (as seen by the confidence interval) is high despite the estimate being significantly different from the actual R_t_. In all experiments, the time when the estimated R_t_ crosses the R_t_ = 1 line corresponds well with the time when the actual R_t_ crosses the same line. Thus, our simulations suggest that R_t_ estimated from surveillance data provides a robust estimate of the time when R_t_ falls below one, which corresponds to the decay phase of the epidemic wave. In experiments with low number of daily tests and when contact tracing is absent, estimated R_t_ rises above one again at the end of the epidemic even though the actual R_t_ remains below one. If the uncertainty of the estimated R_t_ is not taken into account, such a situation might be interpreted as the beginning of a new wave and may lead to incorrect forecasts from epidemiological models.

**Figure 6.**
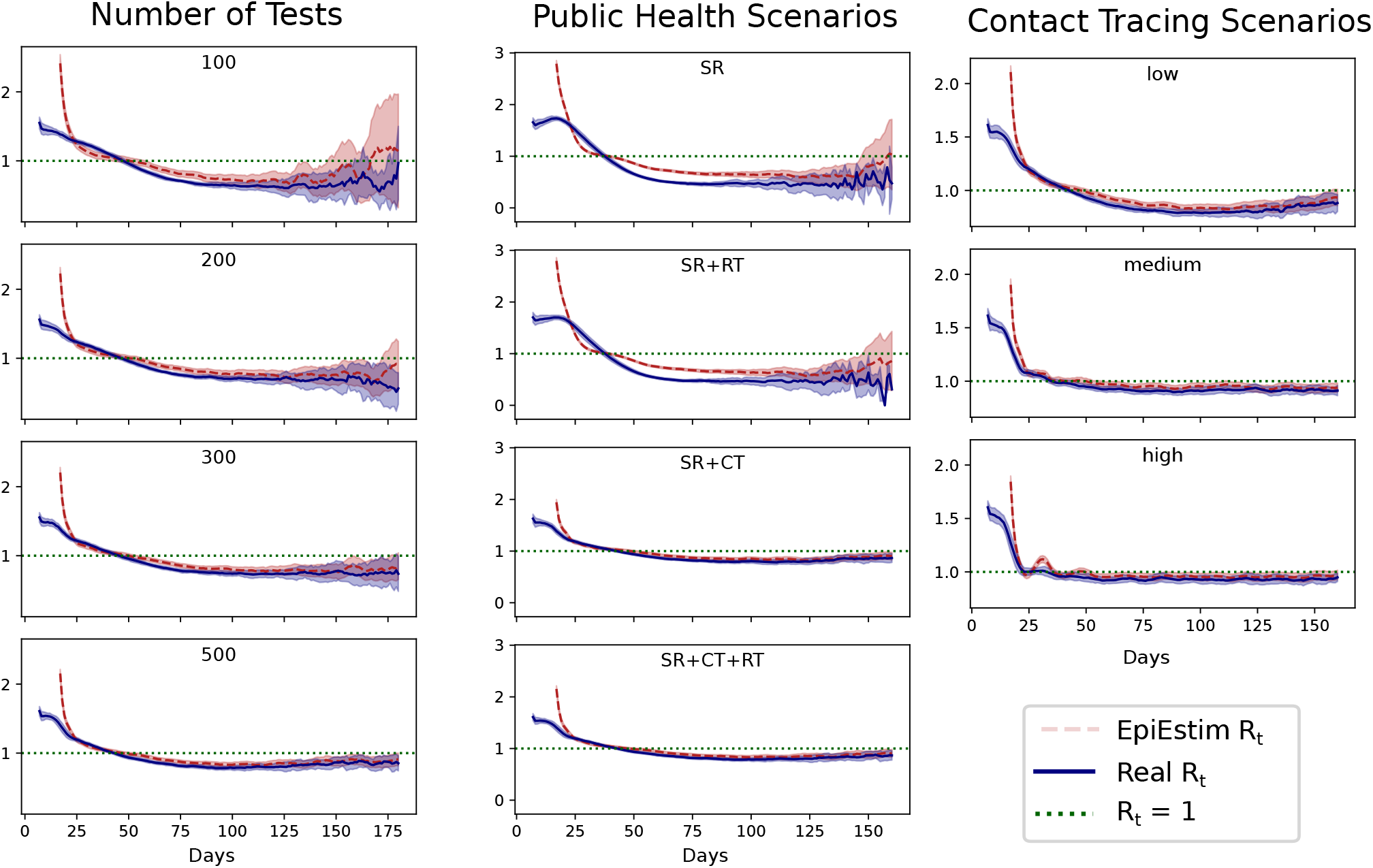
Plot showing the relationship between R_t_ (dashed red line) and the rate of change of true infections per day (solid blue line) for (left) different number of daily tests and (right) different public health scenarios. The shaded red regions around the dashed lines depict the 90% confidence intervals of R_t_. The horizontal grey dotted line indicates R_t_ = 1. The vertical dotted grey line indicates the peak of true infections.

In experiments without contact tracing (SR and SR+RT), estimated and actual R_t_ are in poor agreement over the entire wave except for the time when R_t_ crosses one. In particular, estimated R_t_ is an overestimate in the initial phase, an underestimate near the peak and an overestimate again in the decay phase. On the other hand, experiments with contact tracing provide a more reliable estimate of R_t_ even in extreme scenarios with low number of daily tests and low efficiency of contact tracing(see Figure 6). Thus, our experiments suggest that continuing contact tracing even at low efficiency over the entire course of the epidemic may not only contribute to improved health outcomes, but also to much more reliable estimates of epidemic parameters from the resulting surveillance data.

## 4 Discussion and conclusion

Public health decision making during an epidemic is based on data obtained from surveillance. The sucess of such decisions in controlling health outcomes relies on the fidelity of the surveillance data in representing the actual epidemic, especially when a majority of the cases go unidentified. While metrics derived from surveillance data such as TPR and CFR have been used^14–16^ during the recent COVID-19 epidemic, their actual utility has not been tested in a systematic manner. Thus, the main goal of our work was to use agent-based modelling as a principled approach to both designing and testing such metrics. Put in another way, our work aimed to “construct” the behavior and relationships between different metrics such as TPR and CFR over an epidemic wave and examine how different public health responses lead to different behaviors of such metrics. This constructivist approach allows for a more nuanced use and interpretation of such metrics.

For instance, our results show that regardless of the type of public health response being active, a TPR-threshold based intervention will be activated for too long since it rises much earlier than the number of cases. Thus, a threshold based approach may keep the intervention (such as a lockdown) active for much longer than is necessary to control infection spread, leading to inefficiencies in usage of scarce resources and unnecessary loss of economic output.

The rate of change of TPR peaks earlier than the rate of change of cases, and appears to be a better predictor of when the true number of cases (both detected and undetected) are likely to surge. Since the peak rate of change of TPR occurs just before the epidemic itself peaks, the peak rate of change of TPR provides a way to estimate future requirements for healthcare infrastructure particularly near the epidemic peak. Further, the time when the rate of TPR becomes negative is a good estimate when the number of total cases (not only detected) also begins to decline.

Our results suggest that using metrics such as TPR to compare between cities or administrative wards is problematic since different places may adopt different public health responses and with different efficiencies. Indeed, better contact tracing leads to higher mean TPR in our simulations which may lead to the false impression that the epidemic is more prevalent or that contact tracing efficiency is poor.

Our results also show that CFR tends to peak when the epidemic is already on the decline – even if CFR is assigned based on the date of identification of the infected person rather than date of death – and in some cases has no clear peak at all, making it problematic as a metric for decision making. Furthermore, metrics such as TPR and CFR do not scale linearly with the number of true infections or deaths and thus care must be taken while utilizing TPR or CFR to make inferences about likely health outcomes.

Our results suggest that R_t_ is also a reliable metric in predicting the epidemic peak (i.e, when the total number of cases begins to decline). However, the estimated R_t_ may not provide a good estimate for the real R_t_ at most other times, particularly when contact tracing is not active. EpiEstim provides R_t_ estimates that are too confident, and may lead to false inferences about rate of spread of the infection, particularly before the peak.

One of the main takeaways of our work is the important role contact tracing plays not only in its conventional usage to restrict the rate of infections but also in improving the fidelity of the resulting surveillance data in representing the actual state of the epidemic, in terms of estimating R_t_ and fraction of undetected cases. Thus, contact tracing appears to go beyond its intended role and also acts as an effective “sampling strategy”.

While we present an extensive set of public health scenarios, we do not expect our results to be directly applicable to all real world scenarios. One of the main limitations is that we consider an isolated population with no import/export of infections from external sources. This makes the public health response unusually efficient in identifying infections and leads to very high values of TPR.

Furthermore, we do not account for differential delays in testing, test sensitivity and specificity, reinfections, and transmission through non-airborne medium. We also do not account for pharmaceutical interventions such as medicines, vaccines and antibody therapy and non-pharmaceutical interventions such as lock downs.

Lastly, we assume all our infectious agents are equally likely to transmit the disease. Consequently, our population has no agents who are intrinsically “super-spreaders”. We note, however, that we can nevertheless have super-spreader events that occur due to the network structure of our population, as certain agents have denser contact networks than others.

However, our approach can easily be extended to such situations, and the main aim of this work is to demonstrate the utility of using agent-based modelling to understand the quality of surveillance data that is generated during an epidemic, and not only as a tool for forecasting or evaluation of interventions.

## 5 Acknowledgements

PB acknowledges the Department of Science and Technology, Government of India for the KVPY fellowship. SK acknowledges the Department of Science and Technology, Government of India for the INSPIRE fellowship. PC acknowledges support from Ashoka University and the Mphasis F1 Foundation, and would like to thank Gautam Menon, Vaibhhav Sinha, and Riz Noronha for many useful discussions. The authors acknowledge the support and the resources provided by PARAM Brahma Facility under the National Supercomputing Mission, Government of India at the Indian Institute of Science Education and Research, Pune. The authors thank Ashwini Keskar and Pune Knowledge Cluster for providing the daily epidemiological surveillance data from Pune. The authors would also like to thank the BharatSim team at ThoughtWorks and Ashoka University, with special thanks to Jayanta Kshirsagar and Gaurav Deshkar from the ThoughtWorks Engineering for Research team for many discussions regarding the use and optimisation of BharatSim.

## 6 Additional Information

### 6.2 Data availability statement

Code and simulation data underlying this work have been made available on the GitHub repository https://github.com/SoumilK1/Interpreting-epidemiological-surveillance-data.

### 6.3 Author contributions

JMM conceptualized the study. PB, SK, and PC designed the epidemiological model and developed the code. SK, PB, and JMM analysed the data and wrote the manuscript. All authors reviewed and approved the final version of the manuscript.

## Supporting Information

**S1 Appendix Description of the agent-based network model** We describe the construction of the synthetic population that we used to obtain our simulation results, along with details of the underlying network structure and the schedules of different agents in our model.

**S2 Appendix Details of epidemic simulations** The disease-progression in a single well-mixed compartmental model is briefly described. We also show the correspondence between the infectivity *λ*_*S*_ and the reproductive ratio *R*_0_. Lastly, we provide the epidemic curves for our network model using different values of *λ*_*S*_.

**S3 Appendix Epidemic outcomes of different experiments** We compares the disease trajectories of infections and deaths across multiple intervention scenarios, showing that increased contact tracing and number of tests lead to better epidemic outcomes.

**S4 Appendix Evaluation of CFR as a forecast metric** We provide a comparison of the trajectories of the CFR for different values of true infections across multiple interventions, showing that its utility as a forecast metric of the epidemic peak is limited.

**S5 Appendix Analysis of public health data from Pune** We provide a brief statistical analysis of the contact tracing and testing data from Pune, and compare the TPR and the number of positive cases in Pune city as a function of time.

**S6 Appendix Sensitivity to intervention parameters** We perform a sensitivity analysis of the phase relationships between the trajectories of the TPR and true infections, as well as their respective time derivatives, across scenarios varying in quarantine efficacy, daily testing capacity, and contact tracing efficiency.

**S7 Appendix Comparing R**_**t**_ Compares R_t_ obtained from the underlying epidemic and the EpiEstim R_t_ using both true and observed data. Compares the R_t_ from simulation and EpiEstim for scenarios with high daily tests.

**S8 Appendix Distribution of tests** We show the number of tests used as a function of time in different intervention strategies.

