## Supporting Information for "Interpreting epidemiological surveillance data: A modelling study based on Pune City"

### S1 Appendix: Description of the agent-based network model

#### 1.1 Agents

Agents are the fundamental units of any agent-based simulation. In our simulations, the agents represent the people living in a hypothetical ward based on the wards in Pune city. There are 100,000 agents in the population whose ages are demographically distributed using data from a report of the National Centre for Disease Informatics, and Research [1] on the city of Pune. Each agent has a set of attributes that remain fixed throughout the course of the simulated epidemic. As shown in Table S1, these include a unique agent ID, age, essential worker (a value of 1 implies that they are an essential worker and 0 otherwise), a house ID, a workplace ID, a hospital ID, a cemetery ID, and a neighbourhood ID. The agents are modelled to mimic the schedules and behaviours of real people during an epidemic. As the simulation progresses, agents may become infected and transition through various disease states.

| Agent_ID | Age | essential_worker | HouseID | WorkPlaceID | HospitalID | CemeteryID | NeighbourhoodID |
|---|---|---|---|---|---|---|---|
| 1 | 13 | 0 | 1008 | 619 | 1 | 1 | 11 |
| 2 | 41 | 0 | 10198 | 2084 | 3 | 1 | 100 |
| 3 | 57 | 0 | 17 | 1183 | 5 | 1 | 1 |
| 4 | 58 | 0 | 19844 | 2236 | 1 | 1 | 198 |
| 5 | 38 | 0 | 1349 | 1805 | 5 | 1 | 15 |
| 6 | 54 | 0 | 17668 | 1601 | 2 | 1 | 175 |
| 7 | 13 | 0 | 20570 | 1078 | 3 | 1 | 205 |
| 8 | 26 | 0 | 17483 | 1452 | 2 | 1 | 173 |
| 9 | 25 | 0 | 9054 | 776 | 5 | 1 | 90 |
| 10 | 20 | 0 | 7927 | 1785 | 4 | 1 | 78 |

Table S1: A Snapshot of the input synthetic population file.

#### 1.2 Locations

The model network structure contains geographical locations which are classified as – House, Neighbourhood, Office, Hospital or Cemetery. The network represents the layout of a typical ward in the city of Pune. People are uniformly distributed amongst 25000 houses, with each house containing 4 people on average. While creating the synthetic population, each agent is randomly assigned a house ID based on a uniform prior. The houses themselves are log-normally distributed in 250 neighbourhoods, with each neighbourhood having 100 houses on average, so each neighbourhood includes 400 people on average. A person is assigned a neighbourhood ID based on the neighbourhood in which their house is located, and thus, neighbourhood IDs are not assigned randomly. There are 2500 offices in the network, and each office has 40 employees on average. An office refers to a general area of work/study, such as a workplace cabin/classroom. The office IDs are assigned randomly to people between the ages of 5 and 60. There are 5 hospitals in the network, and hospital IDs are randomly assigned to every person. For essential workers, these hospital IDs designate their workplaces, but for the rest of the population, they simply denote the location of hospitalized agents. There is a single cemetery where all the people are buried after death. The purpose of this cemetery is to remove 'dead' agents from the simulation.

#### 1.3 Schedules

One step of the simulation, denoted by a tick is equivalent to 4 hours and thus one day comprises 6 ticks. All people in the simulation follow certain pre-defined daily schedules according to their designation. People between the ages of 5 and 60 are 'employees', and they follow the `Employee schedule`. Note that the use of the term employee covers a broad variety of designations and is not restricted to just office workers. For example, children going to schools follow the same schedule as 'employees' and thus it is redundant to define schools as a separate location or children as students. They come under the same label as Office

| Schedule | Tick 1 | Tick 2 | Tick 3 | Tick 4 | Tick 5 | Tick 6 |
|---|---|---|---|---|---|---|
| `Employee schedule` | House | House | House | Office | Office | Neighbourhood |
| `HC Worker schedule` | House | House | House | Hospital | Hospital | Neighbourhood |
| `non-Employee schedule` | House | House | House | House | House | Neighbourhood |
| `Hospitalized schedule` | Hospital | Hospital | Hospital | Hospital | Hospital | Hospital |
| `Isolation schedule` | House | House | House | House | House | House |

Table S2: The different schedules followed by different people depending on their designation. The first column describes the type of the schedule while the subsequent columns describe the location of every person during the corresponding tick.

and employees respectively. Essential or health care workers (around 0.2% of the population) follow the `HC Worker schedule`. People below the age of 5 or above the age of 60 are non-employees and they follow the `non-Employee schedule`. People who are hospitalized remain in the hospital until they recover or die and hence follow the `Hospitalized schedule`. The `Isolation schedule` is followed by `-` people who are quarantined (for 14 days) after testing positive, people who are identified as low risk contacts (and are quarantined for 7 days) and symptomatic people who are isolated before getting tested.

### S2 Appendix: Details of epidemic simulations

#### 2.1 Description of the underlying compartmental model

Agents in our simulations can exist in one of eight disease states, (S)usceptible, (A)symptomatic, (P)resymptomatic, Mildly Infected (MI), Severely Infected (SI), (H)ospitalized, and (R)ecovered. In a well-mixed model with no network structure, the number of individuals of age-group $i$ in each of these compartments at any given time is modelled through a set of coupled differential equations:

$$\frac{\mathrm{d}S_i}{\mathrm{d}t} = -\frac{\lambda_S}{N} S(A + P + MI + SI + H) \tag{1}$$

$$\frac{\mathrm{d}A_i}{\mathrm{d}t} = \gamma_i \frac{\lambda_S}{N} S(A + P + MI + SI + H) - \lambda_A A \tag{2}$$

$$\frac{\mathrm{d}P_i}{\mathrm{d}t} = (1 - \gamma_i) \frac{\lambda_S}{N} S(A + P + MI + SI + H) - \lambda_P P \tag{3}$$

$$\frac{\mathrm{d}MI_i}{\mathrm{d}t} = \delta_i \lambda_P P - \lambda_{MI} MI \tag{4}$$

$$\frac{\mathrm{d}SI_i}{\mathrm{d}t} = (1 - \delta_i) \lambda_P P - \lambda_{SI} SI \tag{5}$$

$$\frac{\mathrm{d}H_i}{\mathrm{d}t} = \sigma_i \lambda_{SI} SI - \lambda_H H \tag{6}$$

$$\frac{\mathrm{d}R_i}{\mathrm{d}t} = \lambda_A A + \lambda_{MI} MI + (1 - \sigma_i) \lambda_{SI} SI + (1 - \mu_i) \lambda_H H \tag{7}$$

$$\frac{\mathrm{d}D_i}{\mathrm{d}t} = \mu_i H \tag{8}$$

where $N$ is the total population. For our model which has an inbuilt network structure, these equations describe the rate of transition at every step and at every location in the network.

#### 2.2 Calibrating the parameter $\lambda_S$

We calibrate $\lambda_S$ to the basic reproduction number $\mathrm{R}_0$ in the absence of public health interventions. We do this by simulating the epidemic for 21 days using a range of $\lambda_S$ values. For each case, we calculate $\mathrm{R}_0$ as the average number of secondary infections of individuals infected on or before day 14. The dependence of $\mathrm{R}_0$ on $\lambda_S$ is shown in Figure S1.

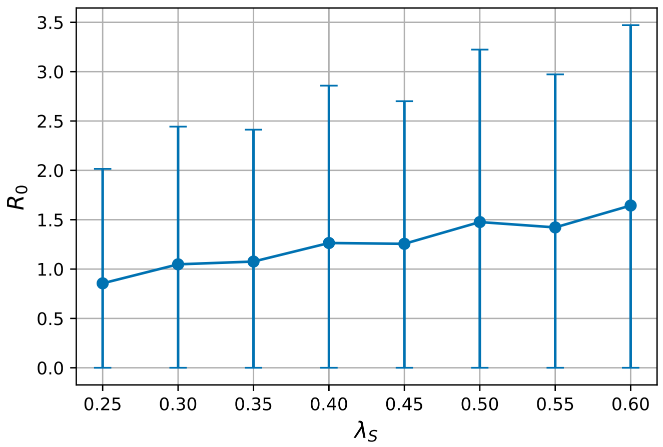

Figure S1: The basic reproductive ratio $\mathrm{R}_0$ obtained from our simulations as a function of $\lambda_S$. The error bars correspond to 1 standard deviation across 30 simulation runs.

##### 2.3 Choosing the parameter $\lambda_S$

Figure S2 shows the number of people in each disease state as the epidemic progresses for different chosen values of $\lambda_S$ in our agent-based model. It is evident that as $\lambda_S$ increases, the peak of the infection curve also increases for all the disease states, but the epidemic lasts for a shorter duration. We choose to run our simulations for $\lambda_S = 0.5$, since it results in about 70% of the population being infected by the end of the epidemic in the case of no interventions. Our calibration results indicate that $\lambda_S = 0.5$ corresponds to an $R_0$ of 1.5.

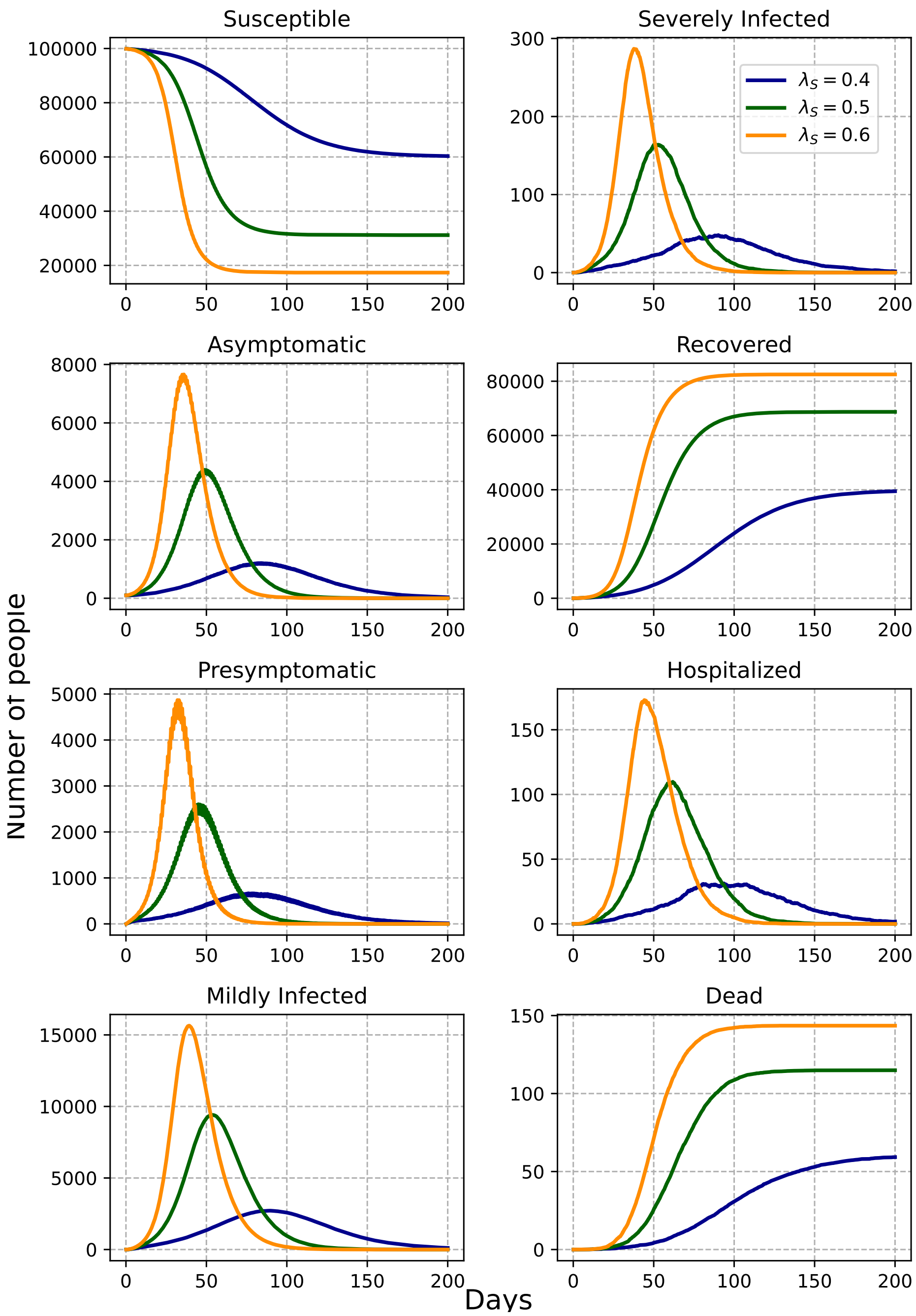

Figure S2: Number of people in different diseases states as epidemic progresses for different values of $\lambda_S$.

### S3 Appendix: Epidemic outcomes of different experiments

#### 3.1 Effect of varying public health response

Figure S3 shows the evolution of the number of true infections and the number of cumulative deaths for different public health scenarios given a fixed number of 500 daily tests. Contact tracing reduces the spread of the epidemic as the SR + CT and the SR + RT + CT scenarios show a significant reduction in the number of infections and the number of cumulative deaths. Random testing, on the other hand, has a negligible effect.

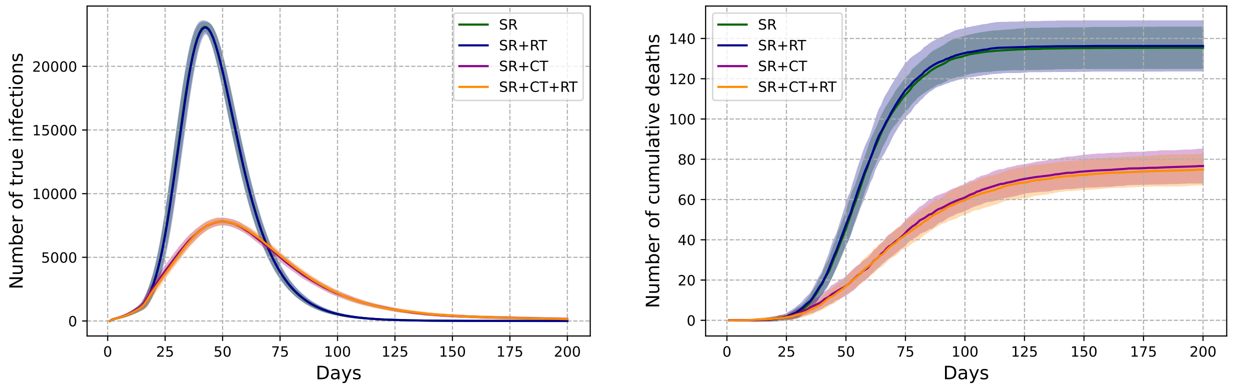

Figure S3: The evolution of the number of true infections (left) and the number of cumulative deaths (right) for different public health response scenarios given a fixed number of 500 daily tests

#### 3.2 Effect of varying the number of daily tests

Figure S4 shows the evolution of the number of true infections and the number of cumulative deaths for different numbers of daily tests given a fixed public health response - SR + CT + RT. An increase in the number of daily tests leads to a decrease in the infection peak and also in the number of cumulative deaths. However, this trend does not continue in a linear fashion as with a subsequent increase in the number of daily tests, the number of true infections and the number of cumulative deaths saturate. The reduction in the infection peak corroborates with results described in other studies [2, 3, 4] which also show that increasing the number of daily tests reduces the spread of the epidemic.

#### 3.3 Effect of varying the extent of contact tracing

Figure S5 shows the evolution of the number of true infections and the number of cumulative deaths for different contact tracing efficiencies given a fixed public health response - SR + CT and a fixed number of 500 daily tests. An increase in the efficiency of contact tracing decreases the spread of the epidemic, reducing both, the number of true infections and the number of cumulative deaths . Similar results have been shown in other studies [5, 6, 7, 8, 9] where increasing the efficiency of digital and/or manual contact tracing led to a reduction in the spread of the epidemic. However increasing the efficiency of contact tracing prolongs the duration of the epidemic wave.

#### 3.4 Comparing epidemic outcomes from all the experiments

In Figure S6, we provide a comparison of epidemic outcomes and public health metrics for all experiments.

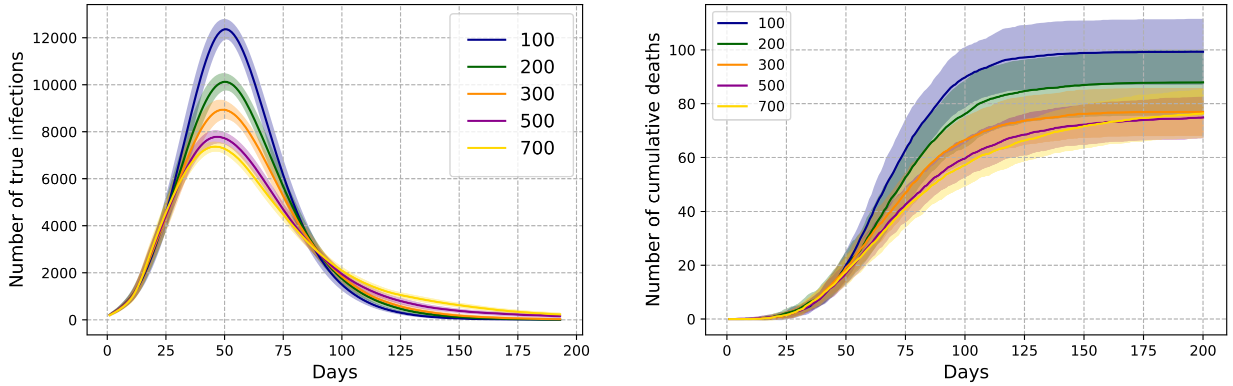

Figure S4: The evolution of the number of true infections (left) and number of cumulative deaths (right), for different number of available daily tests as shown in the legend, given a fixed public health response - SR + CT + RT. All plots correspond to a 7-day rolling average.

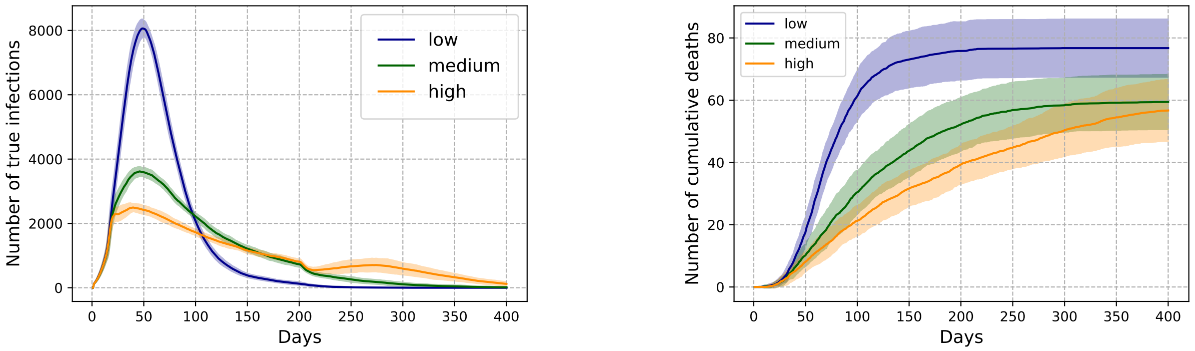

Figure S5: Evolution of the number of true infections (left) and the number of cumulative deaths (right) for different efficiencies of contact tracing, given a fixed public health response - SR + CT and a fixed number of 500 daily tests.

| **Scenario** | **Public health response** | **No.of daily tests** | $\mathbf{f_O}$ | $\mathbf{f_N}$ |
|---|---|---|---|---|
| S1 | SR | 500 | 0.1 | 0.02 |
| S2 | SR + CT | 500 | 0.1 | 0.02 |
| S3 | SR + CT + RT | 300 | 0.1 | 0.02 |
| S4 | SR + CT + RT | 500 | 0.1 | 0.02 |
| S5 | SR + CT + RT | 700 | 0.1 | 0.02 |
| S6 | SR + CT | 500 | 0.05 | 0.025 |
| S7 | SR + CT | 500 | 0.25 | 0.125 |

Table S3: A table describing the parameters for all the scenarios given in Figure S6

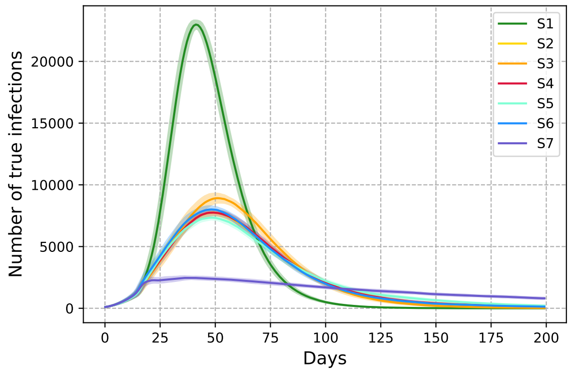

(a) Number of true infections

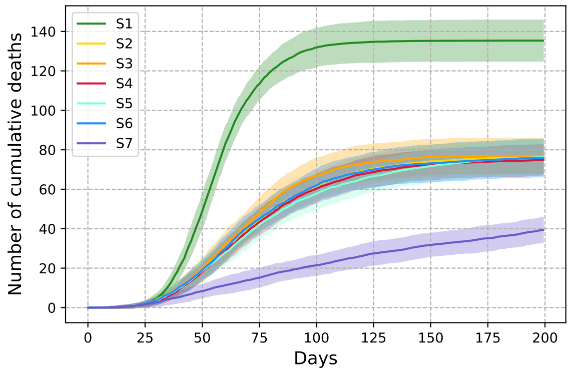

(b) Cumulative number of deaths

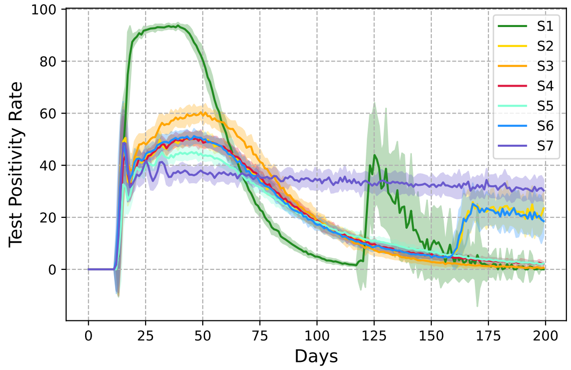

(c) Test Positivity Rate (TPR)

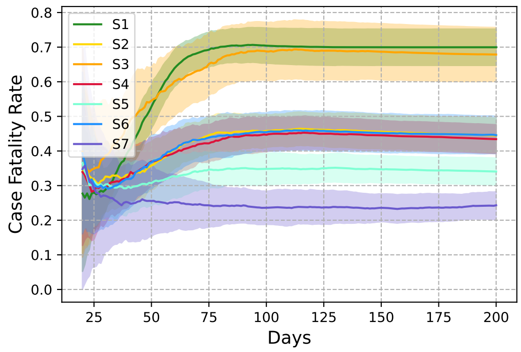

(d) Case Fatality Rate (CFR)

Figure S6: A comparison of the evolution of the (a) number of true infections, (b) cumulative number of deaths, (c) Test Positivity Rate, and (d) Case Fatality Rate for 7 different combinations of the public health response, number of available daily tests, and the efficiency of contact tracing. The values corresponding to each of these are given in Table S3.

#### S4 Appendix: Evaluation of CFR as a forecast metric

Figure S7 compares the evolution of the number of true infections and the CFR. Irrespective of the public health response, the peak of the number of true infections always occurs before the peak of CFR. This implies that CFR is not useful at all in forecasting the peak of the infection curve.

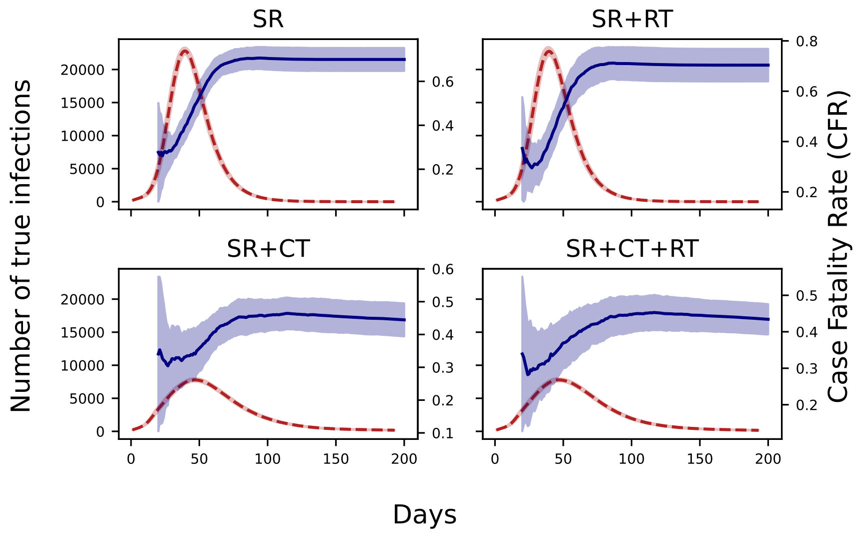

Figure S7: A comparison of the evolution of the number of true infections and CFR for different public health response scenarios given a fixed number of 500 daily tests.

Figure S8 compares the evolution of the number of true infections and the CFR. As shown before, the peak of the number of true infections always occurs before the peak of CFR and thus CFR cannot be used to forecast the peak of the infection.

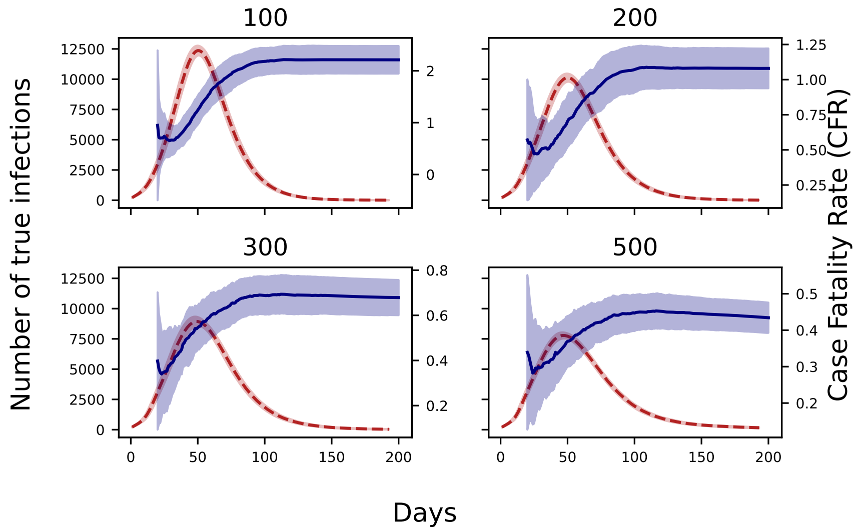

Figure S8: A comparison of the evolution of the number of true infections and CFR for different numbers of daily tests given a fixed public health response - SR + RT + CT.

### S5 Appendix: Analysis of public health data from Pune

#### 5.1 Analysis of contact tracing data

We analysed contact tracing data of the Pune Municipal corporation that was curated in an earlier study[10]. The data contains information about contact tracing in the first wave of the COVID-19 pandemic. The results of the statistical analysis performed on each contact of the dataset is given below in Table S4

| | |
|---|---|
| Data Points | 6271 |
| mean | 7.159783 |
| std | 7.164171 |
| min | 0 |
| 25% | 4 |
| 50% | 5 |
| 75% | 8 |
| max | 71 |

Table S4: Summary of the statistical analysis performed on the contact tracing data

#### 5.2 Comparing Test Positivity Rates and Daily Positive Tests from Pune

We plotted the TPR and Daily Positive Tests from Pune city during the second wave of the COVID-19 epidemic from a dataset curated by the Pune Knowledge Cluster. The peak of TPR appears before the peak of the daily positive cases. The plot is shown in Figure S9.

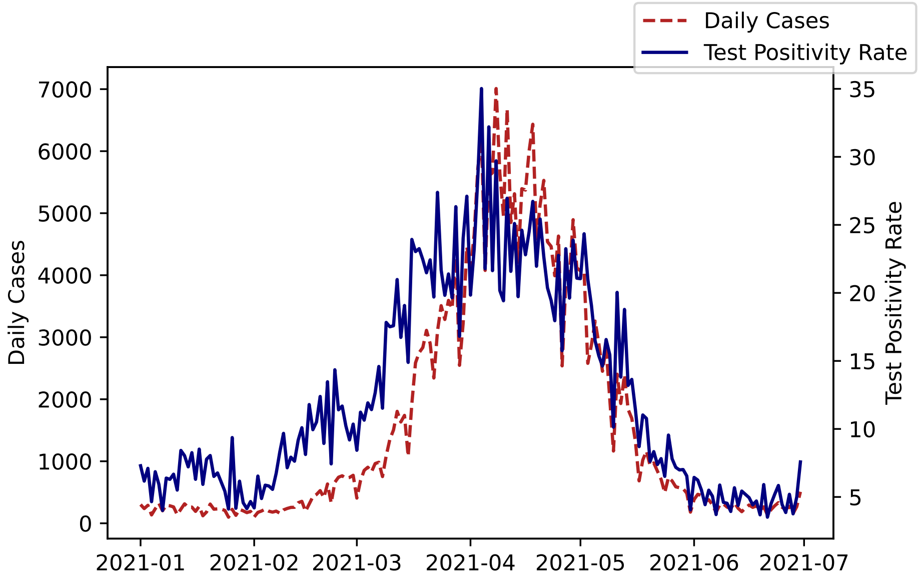

Figure S9: Plot showing the relationship between the Test Positivity Rate and the Number of Positive cases in Pune

### S6 Appendix: Sensitivity to intervention parameters

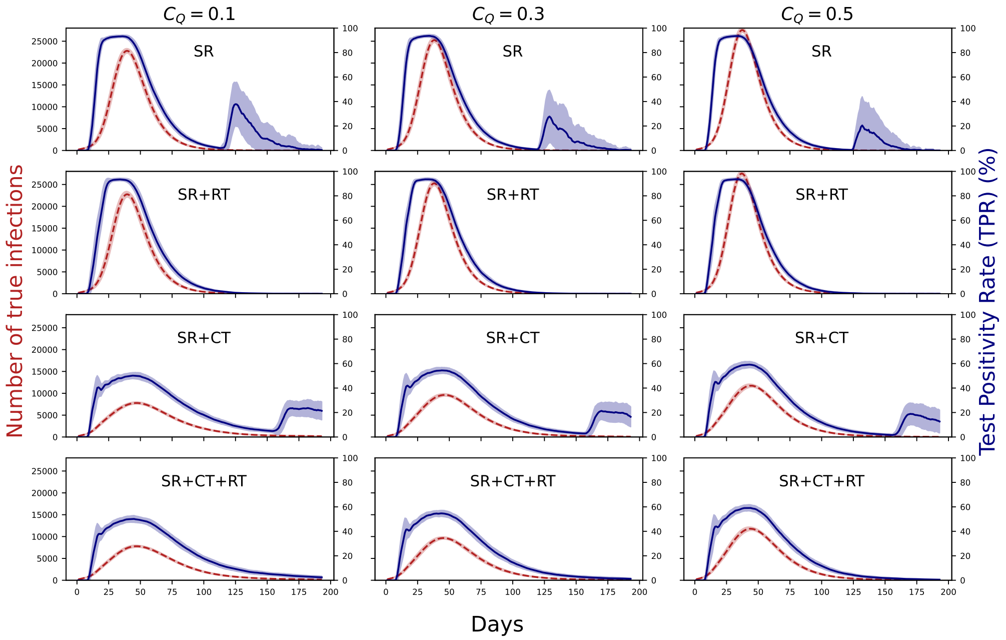

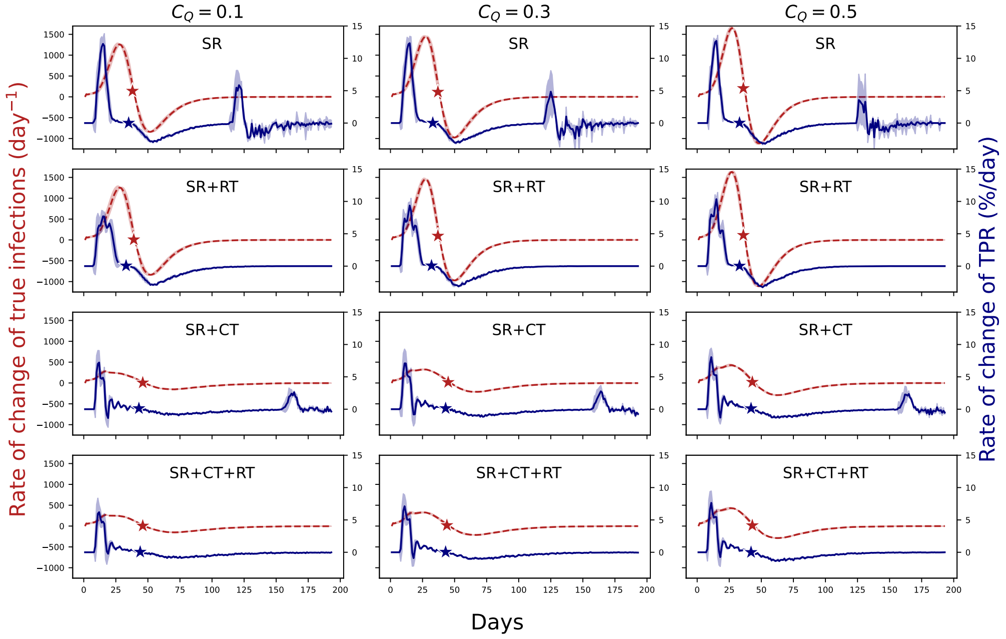

Figure S10: The top figure compares the number of true infections (dashed red line) and the TPR (solid blue line), and the bottom figure compares the rate of change of the number of true infections (dashed red line) with the rate of change of TPR (solid blue line) for different public health response scenarios. The stars denote the days on which the corresponding rates go to 0 (i.e. the days on which the infection and the TPR curves peak). Each column shows results for a different value of $C_Q$ which is a measure of the quarantine efficacy. $C_Q = 0.1, 0.3$, and $0.5$ imply infectivity reductions of 90%, 70% and 50% respectively. All the plots correspond to a 7-day rolling average.

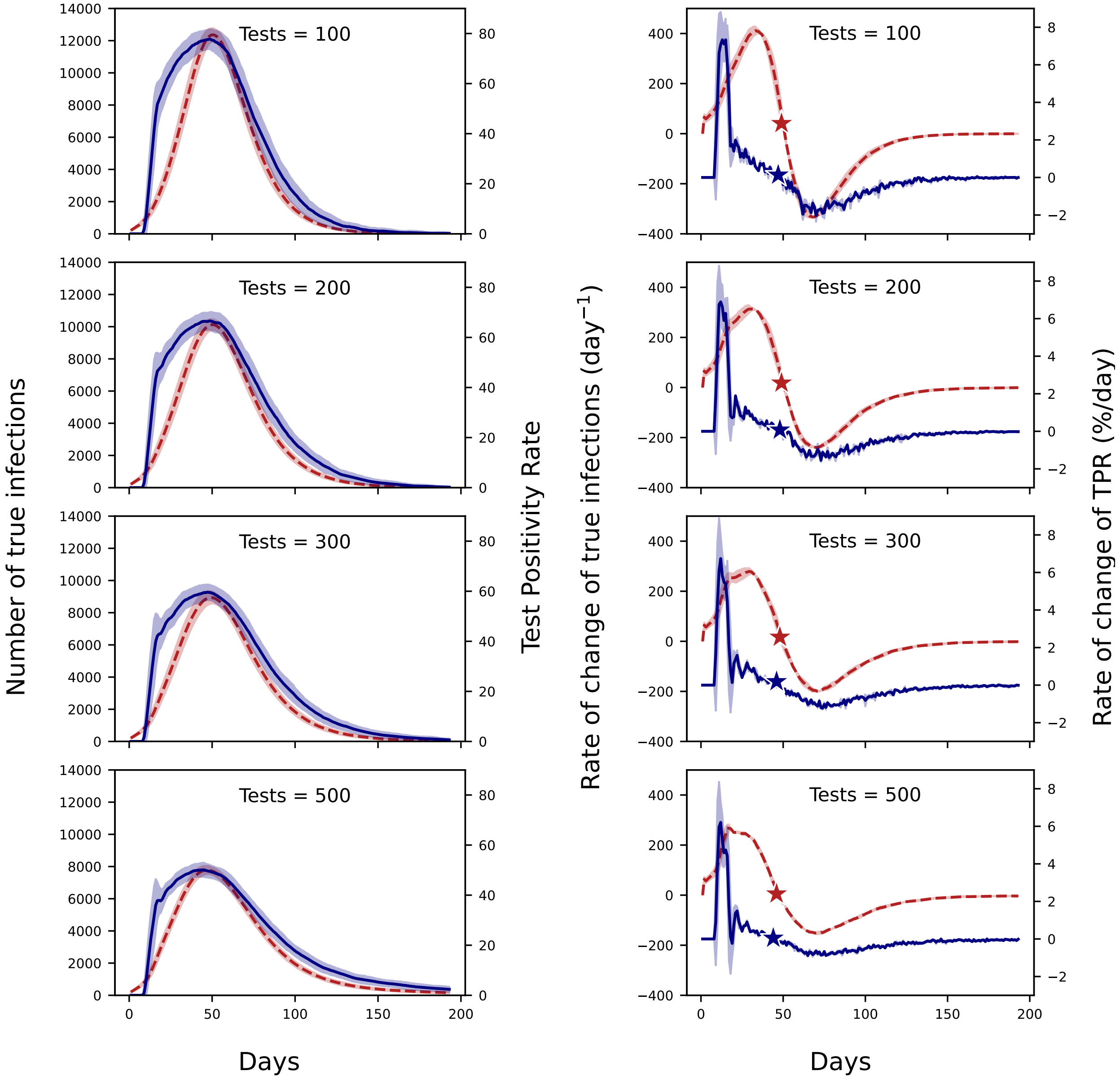

Figure S11: The left panels show a comparison between the number of true infections (dashed red line) and the TPR (solid blue line) and the right panels compare the rates of change of the number of true infections (dashed red line) with the rate of change of TPR (solid blue line) for different numbers of daily tests given a fixed public health response – SR + RT + CT. The stars denote the days on which the corresponding rates go to 0 or the days on which the infection and the TPR curves peak. All the plots correspond to a 7-day rolling average.

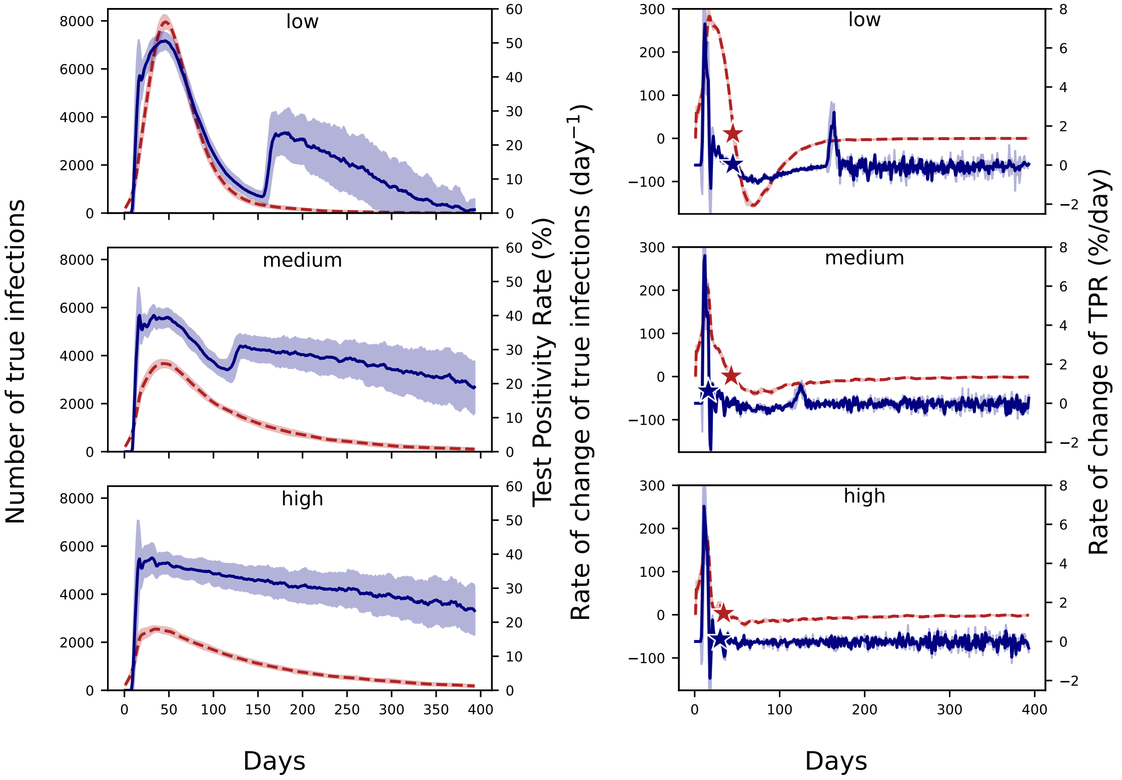

Figure S12: The left panels show a comparison between the number of true infections given (dashed red line) and the TPR (solid blue line) while the right panels compare the rate of change of number of true infections (dashed red line) with the rate of change of TPR (solid blue line) for different efficiencies of contact response keeping a fixed public health response - SR + CT and a fixed number of 500 daily tests. The stars denote the days on which the infection and the TPR curves peak or the days when the corresponding rates go to 0. All the plots correspond to a 7-day rolling average.

### S7 Appendix: Comparing $R_t$

#### 7.1 Parameters used to calculate $R_t$ using EpiEstim

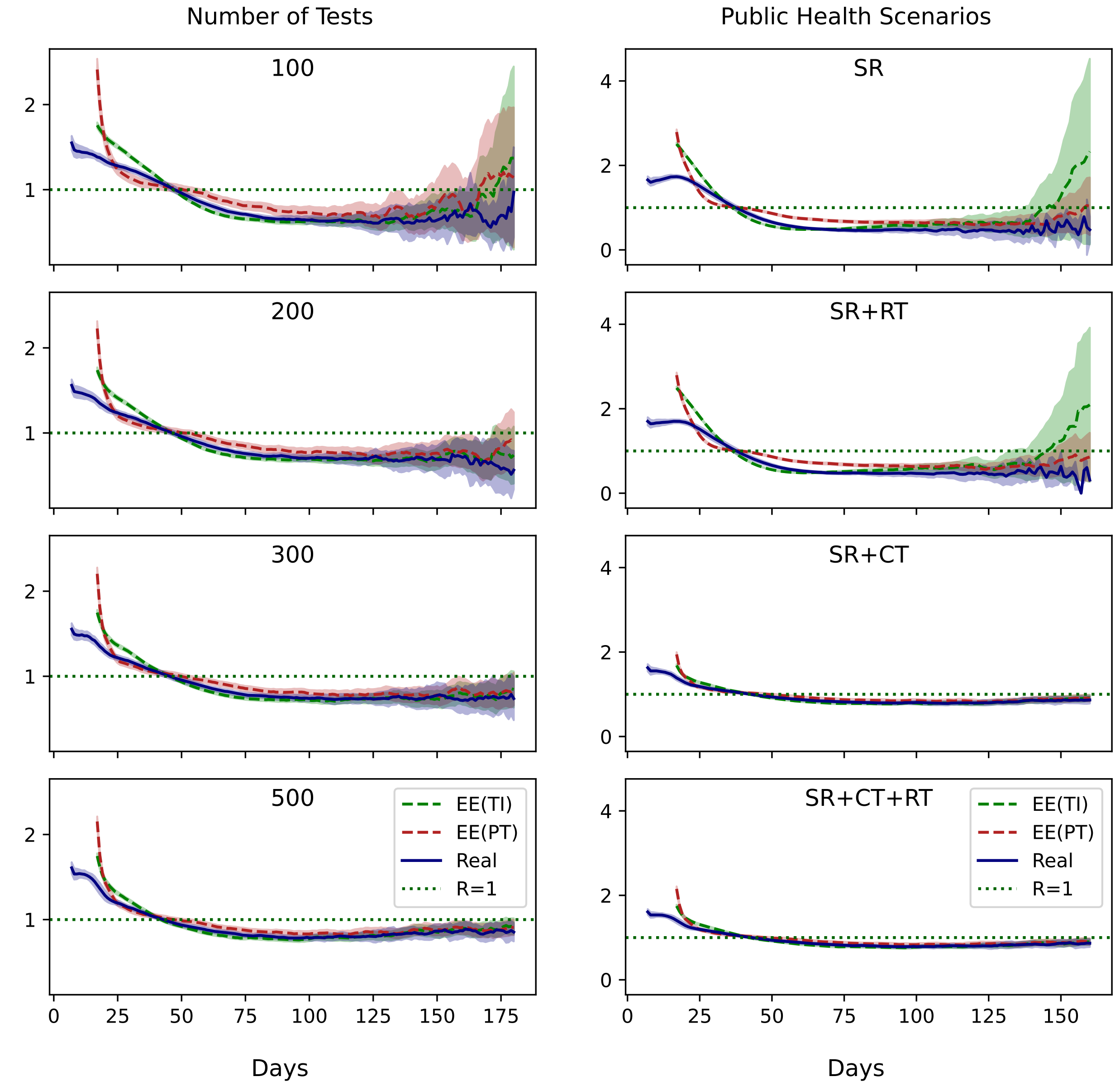

Figure S13: Comparison of the EpiEstim $R_t$ calculated through true infection data (EE(TI)) with EpiEstim $R_t$ (EE(PT)) and the real $R_t$ for different experiments.

#### 7.2 Comparing $R_t$ from true infections and EpiEstim

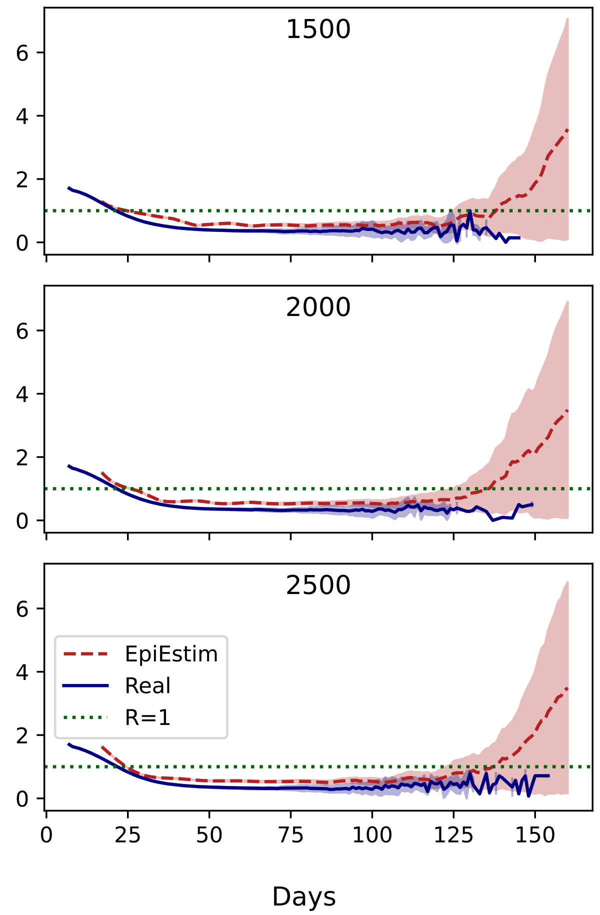

Figure S14: Plot showing the relationship between simulation $R_t$ (dashed red line) and the EpiEstim $R_t$ (solid blue line) for high daily test scenarios. The shaded red regions around the dashed lines depict the 90% confidence intervals of $R_t$

### S8 Appendix: Distribution of tests

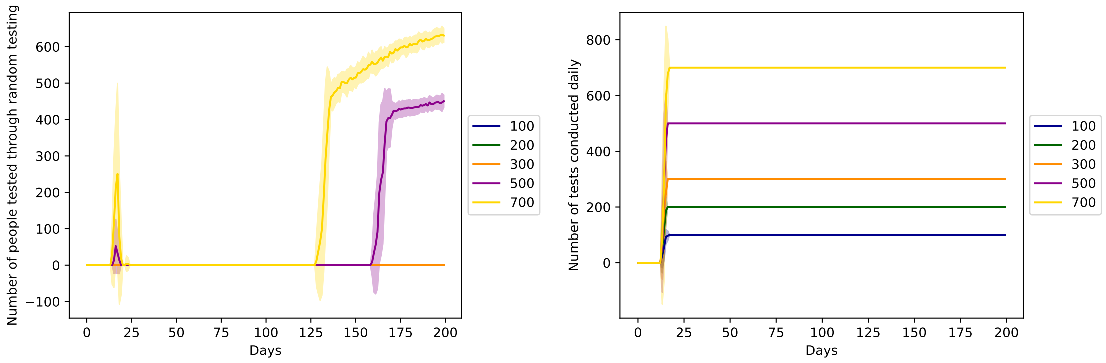

Figure S15: (left) The number of individuals who underwent random testing across different numbers of daily test scenarios. (right) The number of tests used up daily across different numbers of daily test scenarios.

#### 8.1 Proportion of tests used when excess tests are available

We plotted the proportion of tests utilised when excess number of tests are available daily, namely 1500, 2000 and 2500 daily tests. With increasing number of tests available, lesser proportion of tests are utilised.

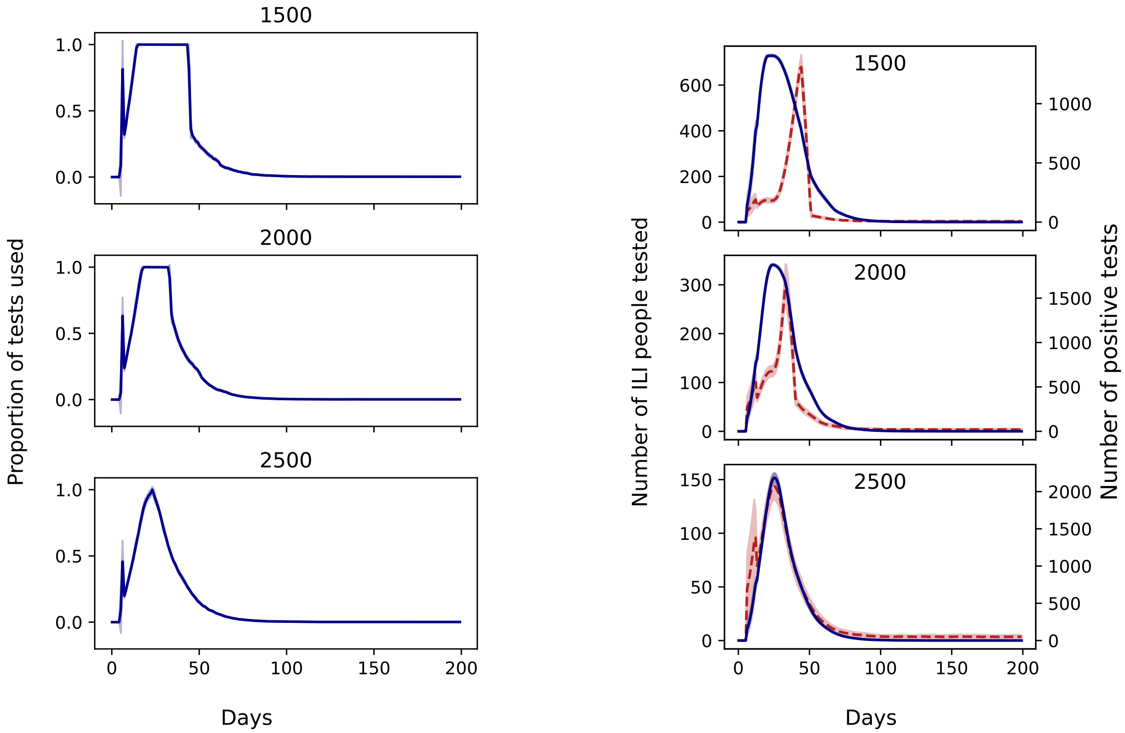

Figure S16: (left) Proportion of tests used daily when excess tests are available for 1500, 2000 and 2500 daily tests. (right) Evolution of the number of positive tests (solid blue line) vs the number of people with influenza-like illnesses (ILI) who are tested (dashed red line) for scenarios with excessive tests.
